# The human milk microbiome varies by environmental factors and is associated with infant growth: findings from the IMiC Consortium

**DOI:** 10.1101/2025.09.18.25336021

**Authors:** Melissa B. Manus, Kelsey Fehr, Chi-Hung Shu, Andrew Mertens, Mark D. DeBoer, Joann M. McDermid, Estomih Mduma, Carl Lachat, Trenton Dailey-Chwalibóg, Laeticia Celine Toe, Lishi Deng, Fyezah Jehan, Muhammad Imran Nisar, Ameer Muhammad, Aneela Pasha, Naveed Iqbal, Waqasuddin Khan, Muhammad Farrukh Qazi, Nima Aghaeepour, Liat Shenhav, Meghan B. Azad

**Affiliations:** Department of Anthropology, University of Texas at San Antonio, TX, USA; Manitoba Interdisciplinary Lactation Centre (MILC), Children’s Hospital Research Institute of Manitoba & Department of Pediatrics and Child Health, University of Manitoba, Winnipeg, Canada; Division of Obstetric Anesthesia, Department of Anesthesiology, Perioperative, and Pain Medicine, Stanford University, Stanford, CA, USA; Department of Biomedical Data Science, Stanford University, Stanford, CA, USA; Department of Pediatrics, Stanford University, Stanford, CA, USA; Division of Epidemiology & Biostatistics, University of California, Berkeley, Berkeley, CA, USA; Division of Pediatric Endocrinology, University of Virginia, Charlottesville, VA, USA; Consultant, Charlottesville, VA, USA; Center for Global Health, Haydom Lutheran Hospital, Tanzania; Department of Food Technology, Safety and Health, Faculty of Bioscience Engineering, Ghent University, Ghent, Belgium; Agence de Formation de Recherche et d’Expertise en Santé pour l’Afrique (AFRICSanté), Bobo-Dioulasso, Burkina Faso; Unité de Nutrition et Maladies Métaboliques; Institut de Recherche en Sciences de la Santé, Bobo-Dioulasso, Burkina Faso; Department of Pediatrics and Child Health, Aga Khan University, Karachi, Pakistan; Vital Pakistan Trust (VPT), Pakistan; Faculty of Life Sciences, BUITEMS, Pakistan; Institute for Systems Genetics, Department of Microbiology, New York Grossman School of Medicine, New York, NY, USA; Department of Computer Science, New York University, New York, NY, USA

## Abstract

Human milk (HM) is a complex ecological matrix that connects mothers and infants to the surrounding environment, and promotes infant growth and health. While certain components of HM are well studied, including its macronutrient content and immune properties, the microbial composition of HM (i.e. the microbiome) is poorly characterized and its impact on infant health phenotypes is largely unknown. We hypothesized that the HM microbiome varies by environmental factors and is associated with differences in growth outcomes among HM-fed infants in settings with elevated rates of undernutrition and growth faltering. We leveraged a large dataset of HM samples (N=451) collected from mothers living in rural Tanzania, rural Burkina Faso, and peri-urban Pakistan around 1 month postpartum as a part of the International Milk Composition (IMiC) Consortium. 16S rRNA bacterial gene sequencing revealed geographic and seasonal signatures of the HM microbiome. Machine learning models identified *Corynebacterium* as a key feature that predicted infant birth season and growth outcomes in each of the three populations, though individual predictive taxa within the genus differed across the models. This study highlights the evolutionary importance of the HM microbiome as a biological system that embeds local environments and is associated with growth phenotypes critical to infant health and survival.

## Introduction

Human milk (HM) is a complex ecological matrix that connects nursing dyads to the surrounding environment and promotes infant health (Christian et al. 2021). HM evolved as an optimal food source for infants that supports early life growth, immune system development, and protection from pathogenic infections (Donovan et al. 2023). HM’s myriad functions are supported by its rich composition, including macro and micronutrients, cytokines, immunoglobulins, and a microbiome—the collection of microbes detectable in HM (Fehr et al. 2020; Moossavi and Azad 2020). The presence of microbes, particularly bacteria, in HM is explained by two non-mutually exclusive ideas: (i) the enteromammary hypothesis, which describes the migration of bacteria from the gastrointestinal tract to the mammary system, likely through assistance from immune cells (Rodríguez 2014), and (ii) the retrograde inoculation hypothesis, where infants transmit bacteria from their skin and oral cavities into HM through the back flow of saliva into mammary ducts during breastfeeding (Ramsay et al. 2004; Moossavi and Azad 2020). These routes emphasize the interconnected nature of the mother-infant-milk triad (Shenhav and Azad 2022).

HM feeding shapes bacterial communities in the infant mouth, gastrointestinal tract, and on the skin (Latuga, Stuebe, and Seed 2014; Fehr et al. 2020; Bogaert et al. 2023). The colonizing bacteria produce essential metabolites like vitamins and short-chain fatty acids (SCFAs) which serve as energy sources for host tissues (den Besten et al. 2013; De Vadder et al. 2014) and promote infant nutrition and growth (Smilowitz et al. 2023). HM also delivers commensal microbes to train and regulate the immune system, contributing to the protective effect of HM feeding against early life illnesses (Oddy 2001). During infection, immune responses divert energy away from linear growth (Urlacher et al. 2018; Garcia et al. 2020), further challenging infant health in settings with elevated risk of early life infection. By supporting microbiome-immune axis development, HM feeding can help infants respond efficiently to infection and mitigate energetic costs, further promoting early life growth.

Since the HM microbiome helps mediate the influence of HM feeding on infant physiology, variation in HM microbial profiles may partly explain differences in growth among HM-fed infants. Like the other bioactive compounds in HM, the microbiome appears sensitive to factors both internal (e.g. nutritional status; infection) and external (e.g. diet; social interactions) to the mother-infant dyad. For example, maternal nutritional status is linked to HM microbial profiles (Daiy et al. 2022) and associated with variation in HM macronutrients and hormones (Adhikari et al. 2022; Sims et al. 2020). Maternal social environments may influence immune factors in HM (Ziomkiewicz et al. 2021) and also shape HM bacterial diversity (Meehan et al. 2018). HM’s sensitivity to the surrounding environment (Ma et al. 2024) is likely adaptive, allowing for the transmission of locally “calibrated” information to the developing infant body (Lackey et al. 2019; Keady et al. 2023; Bornbusch et al. 2024). However, since few studies have cataloged HM variation across populations (Lackey et al. 2019; Meehan et al. 2018), explored its determinants (Ma et al. 2024), or tested for its relationship to infant growth, the evolutionary relevance of the microbiome as a system that connects early life environments to health phenotypes remains elusive.

To address these gaps, we investigated the determinants of the HM microbiome and its connections to infant growth across three geographically and culturally distinct populations in the Global South, which has received less attention in microbiome research compared to cohort-based studies in the Global North (Subbarao et al. 2015; Bisgaard 2004). This work stems from the multi-cohort International Milk Composition (IMiC) Consortium, which aims to characterize global HM profiles and identify associations to infant health (Fehr et al. 2025). In line with these goals, we characterized the microbiome of HM samples collected from mothers living in rural Tanzania, rural Burkina Faso, and peri-urban Pakistan, where undernutrition and growth faltering is of concern. While the larger IMiC Study also includes samples from Canada, we focus on the three IMiC populations where elevated risk of infection that detracts from linear growth may be mitigated by HM feeding during infant microbiome-immune axis development. HM samples were paired with extensive metadata on maternal physiology, physical and social environments, and infant anthropometry from the Early Life Interventions for Childhood Growth and Development in Tanzania (“ELICIT”) trial (DeBoer et al. 2018), the Micronutriments pour la Santé de la Mère et de l’Enfant III (“MISAME-III”) trial in Burkina Faso (Vanslambrouck et al. 2021), and the MUMTA-Lactating Women Trial (“MUMTA”) in Pakistan (Muhammad et al. 2020). The current manuscript addresses three main research questions about the HM microbiome in these contexts:

1. How do HM microbiome profiles differ across three populations in IMiC?
2. What are the determinants of HM microbiome profiles within each IMiC population?
3. Is there a relationship between the HM microbiome and infant growth in IMiC?

## Results

### Participant characteristics differed across populations

We analyzed the microbiome profiles from a subset of HM samples collected around 1 month postpartum (mean (sd) 28.3 (10.2) days, **Table 1**). This included 123, 189 and 139 samples from ELICIT, MISAME-III and MUMTA, respectively. The majority of infants were exclusively HM-fed (99.6%, 449/451). Various maternal, infant and environmental factors differed across the studies (**Table 1**). For instance, parity ranged from a median of two in MUMTA to four in ELICIT. Average maternal BMI was slightly lower in MUMTA (19.4 kg/m^2^) than ELICIT (22.2) or MISAME-III (22.8), and median household size was large overall, ranging from five to ten people. The prevalence of infant stunting at three months (LAZ<-2) was 20% and 19% in MUMTA and ELICIT, and 11% in MISAME-III.

**Table 1.** Study characteristics and population demographics for the three IMiC studies included in the current manuscript.

| Study Characteristics n (%) or mean (sd) | ELICIT<br>NCT03268902 | MISAME-III<br>NCT03533712 | MUMTA<br>NCT03564652 | <u>Overall</u><br><u>IMiC ~ 1</u><br><u>month</u> |
| --- | --- | --- | --- | --- |
| Location | Haydom,<br>Tanzania<br>Rural | Houndé, Burkina<br>Faso<br>Rural | Karachi,<br>Pakistan<br>Urban | - |
| Infant birth years | 2017-18 | 2021 | 2018-19 | 2017-21 |
| Total N milk samples analyzed | 123 | 189 | 139 | 451 |
| **Dyad characteristics** |  |  |  |  |
| Parity, median (IQR) | 4 (2.0-6.0) | 3 (1-4) | 2 (1-4) | 3 (1.0-4.5) |
| Maternal BMI kg/m<sup>2</sup>* | 22.2 (3.6) | 22.8 (3.1) | 19.4 (1.8) | 21.6 (3.3) |
| Maternal age, years | 28.9 (7.0) | 24.2 (5.7) | 24.2 (4.9) | 25.5 (6.2) |
| Infant age (at milk sample collection), days | 28.0 (3.0) | 18.4 (2.5) | 42.0 (1.6) | 28.3 (10.2) |
| Gestational age, days | Unavailable | 279.5 (9.9) | 272.3 (10.7) | 276.4 (10.8) |
| Infant sex (male) | 57 (46.3) | 90 (47.6) | 61 (43.9) | 208 (46.1) |
| Exclusive breastfeeding | 123 (100.0) | 187 (98.9) | 139 (100.0) | 449 (99.6) |
| **Environmental** |  |  |  |  |
| N people living in household, median (IQR) | 7 (5.0-9.0) | 5 (3.0-9.0) | 10 (7.0-13.0) | 7 (5.0-10.8) |
| Improved water source (tap) | 76 (61.8) | 104 (55.0) | 133 (0.95) | 312 (69.2) |
| Improved flooring (non-clay) | Unavailable | 118 (62.4) | 135 (97.1) | 253 (77.1) |
| Birth season, pre-harvest | 42 (34.1) | Not applicable | Not applicable | - |
| Birth season, rainy | 63 (51.2) | 150 (79.4) | 42 (30.2) | 255 (56.5) |
| **Infant growth and health** |  |  |  |  |
| Stunting** at 3 months | 23 (18.7) | 21 (11.3) | 21 (20.0) | 65 (15.7) |
| Stunting** at 6 months | 27 (22.0) | 14 (7.7) | 13 (11.3) | 54 (12.9) |
| Small HCAZ*** at 3 months | 8 (6.6) | 12 (6.5) | 20 (19.0) | 40 (9.7) |
| Small HCAZ*** at 6 months | 2 (1.6) | 13 (7.2) | 20 (17.4) | 35 (8.4) |
| LAZ, at 3 months | -1.0 (1.0) | -0.6 (1.3) | -1.1 (1.2) | -0.9 (1.2) |
| LAZ, at 6 months | -1.1 (1.0) | -0.5 (1.1) | -0.9 (1.1) | -0.8 (1.1) |
| HCAZ at 3 months | -0.3 (1.1) | -0.4 (1.0) | -1.1 (1.0) | -0.5 (1.1) |
| HCAZ at 6 months | -0.3 (0.9) | -0.6 (1.0) | -1.1 (1.0) | -0.6 (1.0) |
| Infant diarrhea^ | 79 (64.2) | 23 (12.2) | 41 (32.3) | 143 (32.6) |
| Infant cough^ | 35 (28.5) | 34 (18.1) | 85 (65.9) | 154 (35.0) |
| Infant vomit^ | Unavailable | 9 (4.8) | 21 (16.5) | 30 (9.5) |

\*mean (sd) days postpartum at BMI measurement: ELICIT, 7.1 (3.4); MISAME-3, 25.3 (11.4); MUMTA, 33.27 (10.01). \*\*Infant HAZ SD < -2; \*\*\*HCAZ SD < -2; BEP, Balanced energy protein supplementation; RCT, randomized controlled trial. Population characteristic values are n(%) or mean ± standard deviation.
^ Symptoms reported by mothers.

### HM bacterial diversity and abundances varied across populations

We detected distinct patterns in the HM microbiome across populations using multiple metrics and statistical approaches (**Table 2**; **Figure 1**). While we observed a “core” set of five HM Amplicon Sequence Variants (ASVs) shared across all samples (*Corynebacterium amycolatum.1*; *Corynebacterium.8*; *Staphylococcus.126*; *Streptococcus.341*; *Streptococcus.38)*, ELICIT HM samples harbored the largest set of unique ASVs (**Figure 1a**) as well as higher bacterial diversity and richness compared to MISAME-III or MUMTA (**Figure 1c**). Bacterial relative abundances also differed across the populations (**Figure 2a**). In MISAME-III, *Priestia.20* and *Bacillus_A.12* showed the largest log-fold increases compared to ELICIT (2.48 and 2.62, respectively). *Bacillus_A.12* (2.20) and *Acinetobacter baumannii* (2.44) were enriched in MISAME-III compared to MUMTA. The largest difference between MUMTA and ELICIT was in *Corynebacterium.17* (2.10 log-fold difference in relative abundance). 26 particularly heterogeneous ASVs were differentially abundant in all three pairwise comparisons (**Figure 2b**). We also identified population-level differences in genus richness (N ASVs within a genus; **Figure 2c**). Richness of *Acinetobacter, Dietzia, Paracoccus*, and *Streptococcus* differed among all populations (p(BH)<0.05). Consistent with overall richness, most genera had higher richness within ELICIT, however *Paracoccus* had higher richness in MUMTA (2.16 ASVs) compared to ELICIT (1.53; p(BH)= 0.009) and MISAME-III (1.12; p(BH)<0.001). *Corynebacterium* had the highest richness within each site and was also significantly higher in ELICIT compared to MISAME-III (p(BH)<0.035).

**Figure 1.**
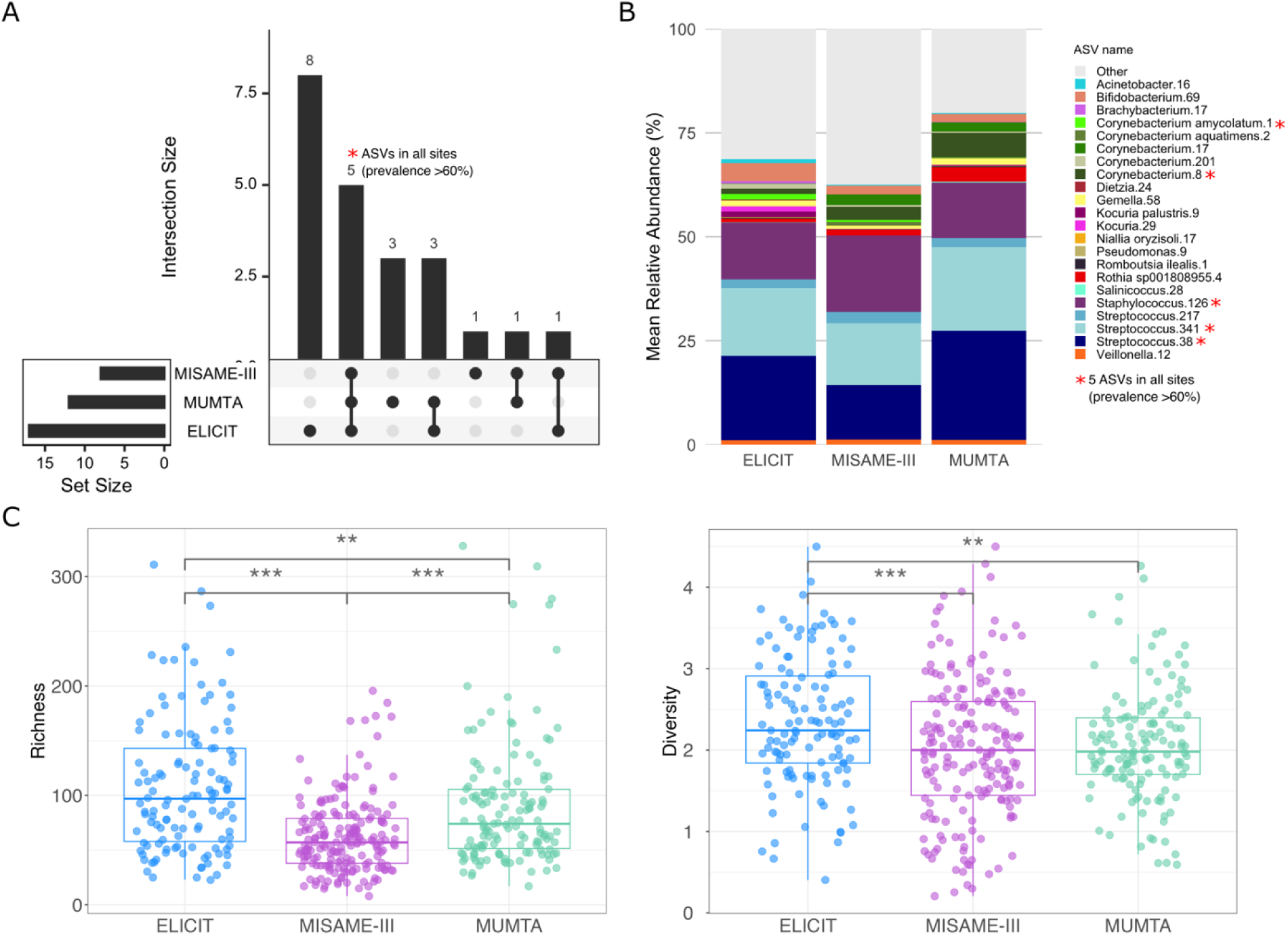
Variation in the milk microbiome across the three IMiC populations. (a) 5 milk ASVs constituted the core microbiota (present in ≥60% of all samples). (b) Relative abundance of ASVs present in ≥60% of milk samples in any population. (c) Shannon diversity and bacterial richness. ***p ≤ 0.001, **p ≤ 0.01, Wilcoxon rank sum test.

**Figure 2.**
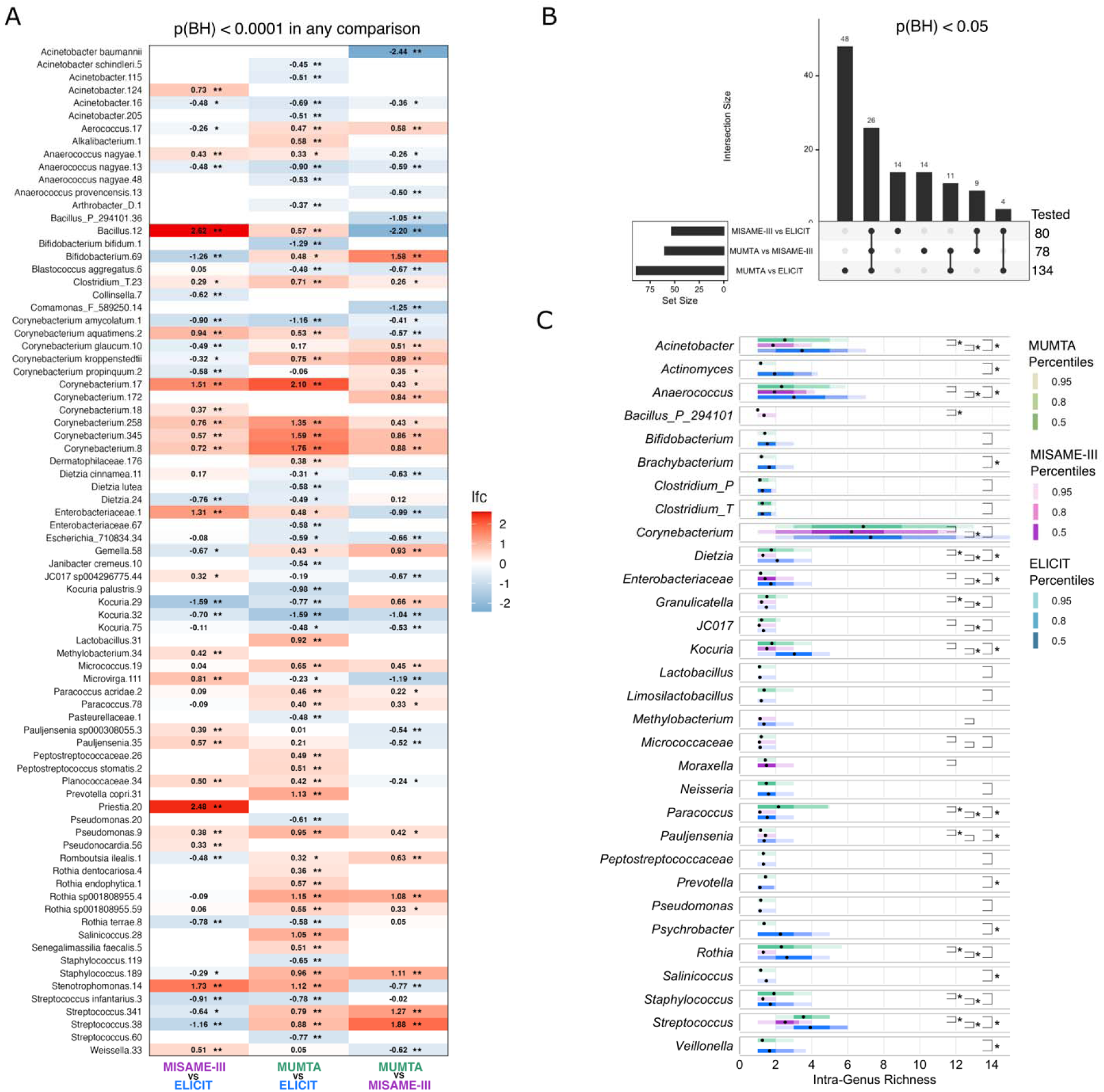
Variation in the milk microbiome ASVs with differential abundance between the studies. (a) Log-fold changes of the most differentially abundant ASVs (p(BH)<0.0001) detected by ANCOM-BC2 in at least one of the three comparisons: 1) MISAME-III vs ELICIT, 2) MUMTA vs ELICIT, and 3) MUMTA vs MISAME-III. Log-fold changes are annotated as *p(BH)<0.05, **p(BH)<0.0001. Empty cells reflect an ASV that was absent from a given comparison (present in <10% of samples in one of the studies being compared). (b) Summary of differentially abundant ASVs across all comparisons. Set size represents the number of ASVs identified as significantly different between studies (p(BH)<0.05, ASVs present in >10% prevalence in both studies were tested). A singular unconnected dot represents the collection of ASVs that differed significantly in a single comparison. For instance, most of the tested ASVs differed significantly only between MUMTA and ELICIT (48 ASVs). Total ASVs tested: 80 (MISAME-III-ELICIT overlap), 78 (MUMTA-MISAME-III overlap), 134 (MUMTA-ELICIT overlap) (c) Intra-genus richness comparisons across sites for any genus present in at least two of the three sites, and including only those samples that contained ≥1 ASV from the genus. Details: Intra-genus sample richness is the number of ASVs from a genus within a given sample. Annotations include the mean (black point), and percentiles (shaded intervals of genus richness that 50, 80 and 95 percent of samples are within).

**Table 2.**
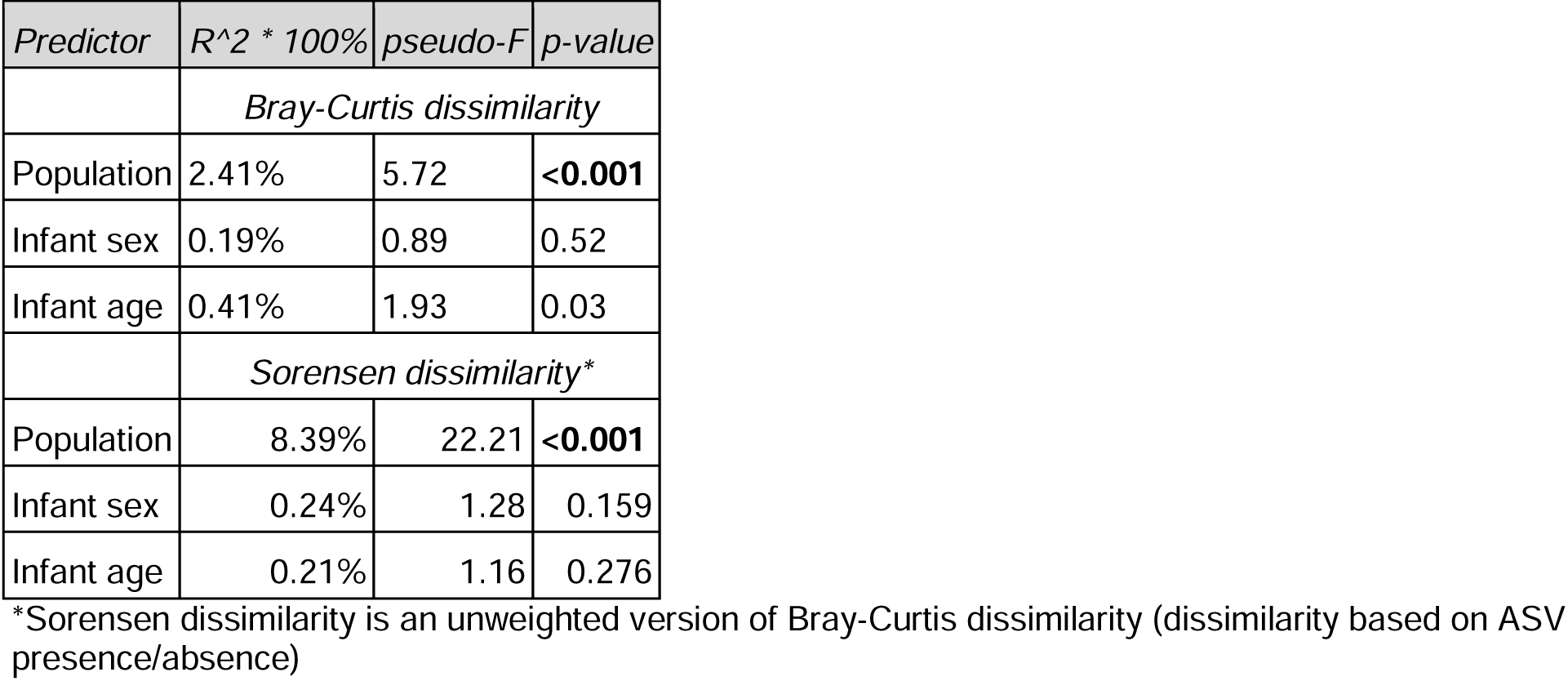
Results of PERMANOVA with bacterial community composition åas the outcome, all samples combined.

| Predictor | R<sup>2</sup> * 100% | pseudo-F | p-value |
| --- | --- | --- | --- |
| *Bray-Curtis dissimilarity* |  |  |  |
| Population | 2.41% | 5.72 | **<0.001** |
| Infant sex | 0.19% | 0.89 | 0.52 |
| Infant age | 0.41% | 1.93 | 0.03 |
| *Sorensen dissimilarity\** |  |  |  |
| Population | 8.39% | 22.21 | **<0.001** |
| Infant sex | 0.24% | 1.28 | 0.159 |
| Infant age | 0.21% | 1.16 | 0.276 |

\*Sorensen dissimilarity is an unweighted version of Bray-Curtis dissimilarity (dissimilarity based on ASV presence/absence)

### Variation in the HM microbiome was explained by environmental factors

Infant birth season was the most significant predictor of HM microbiome profiles across populations (**Figure 3a**; **Table 3; Table S1)**. Categorized as rainy vs. non-rainy, birth season explained considerable variation in HM microbiome profiles in MUMTA (1.7%), and MISAME-III (1.1%) (**Figure 3a**), whereas in ELICIT, the variable explained more variation when categorized as pre-vs. post-harvest (1.2% vs. 0.9%). In ELICIT, maternal age (1.1%) and birth location (1.9%) were also important predictors of HM microbiome profiles, and along with birth season, remained significant predictors in multivariate models of beta diversity (**Table S1**). Improved water source predicted these same outcomes in ELICIT and MISAME-III. In MUMTA, maternal age, parity, and BMI explained significant variation in HM microbiome profiles (1.1%, 1.2%, and 1.1%, respectively), while only birth season significantly predicted beta diversity in multivariate models. Although HM bacterial alpha diversity and richness did not differ by birth season in any population (**Figure 3b**), many ASVs displayed differential abundances based on this variable (**Figure 3c; Table S2-S4**). 87 ASVs differed by birth season in ELICIT (pre-vs. post-harvest), and 58 and 71 in MUMTA and MISAME-III, respectively (rainy vs. non-rainy).

**Figure 3.**
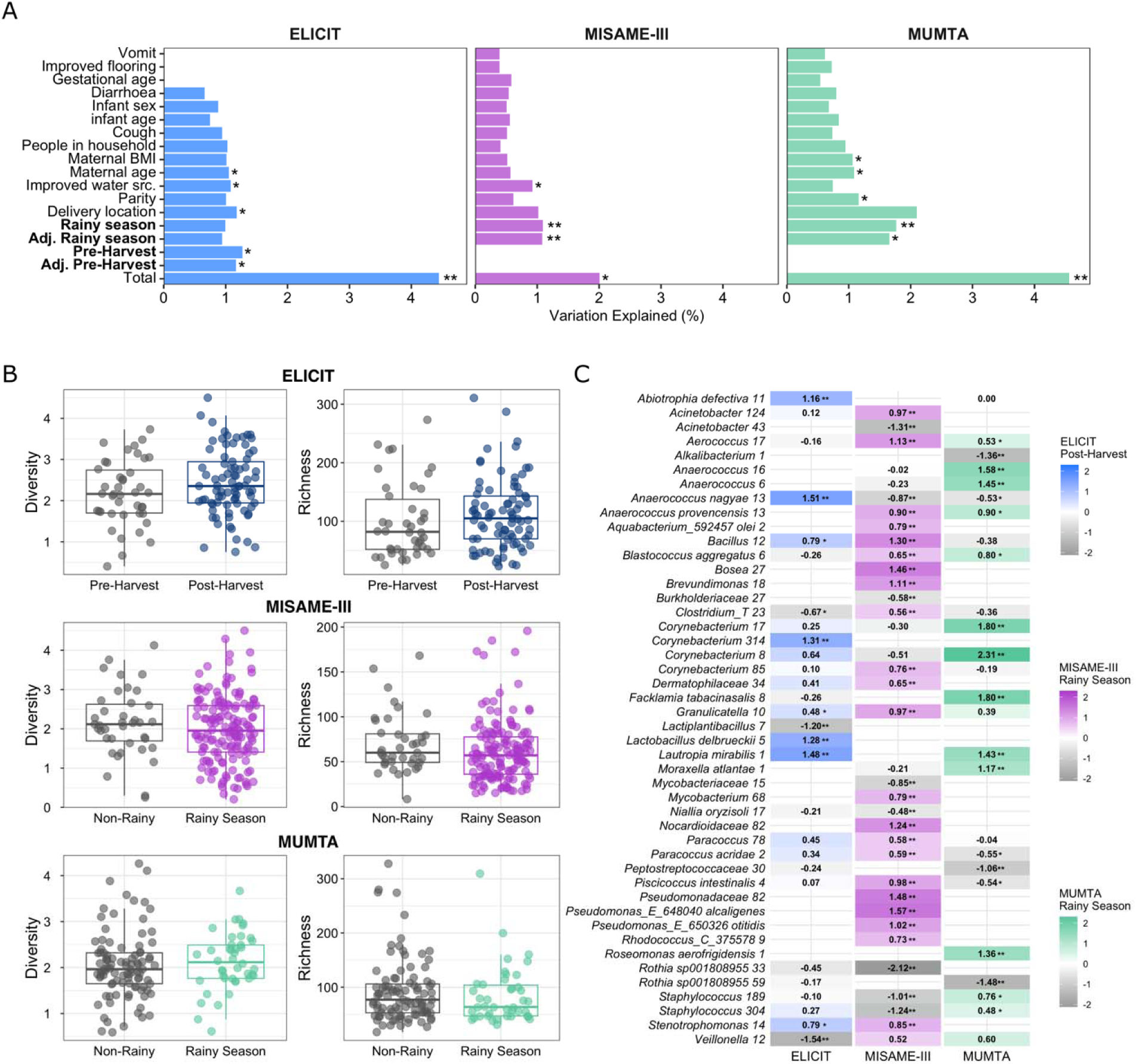
Season of infant birth was the most consistent factor associated with the microbiome. (a) Percent of variation in milk microbiome composition explained by maternal, infant, and environmental factors assessed with redundancy analysis (RDA). Multivariate models (‘Total’, and ‘Adj.’ on the x-axis) include only variables that were significant in univariate RDA. ‘Adj. Pre-Harvest’ and ‘Adj. Rainy season’ represents the variation explained after removal of covariate effects using partial RDA. Variables on the x-axis are ordered from the strongest to weakest explanatory power (averaged across sites). Also see Table S1. *p<0.05, **p<0.001. (b) Alpha diversity (Shannon diversity and richness) displayed minimal variation by infant birth season. All p-values ≥ 0.05, Wilcoxon rank sum tests. (c) Log-fold changes of the most differentially abundant ASVs detected with ANCOM-BC2 (ASVs with p(BH)<0.0001 in any study are shown; blank cells indicate that the ASV was not present in that study with prevalence >10%; delivery in the pre-Harvest season in ELICIT and the non-rainy season in MISAME-III and MUMTA as the referent groups). Also See Table S2, S3 and S4. *p<0.05, **p<0.0001

**Table 3.** Results of PERMANOVA with bacterial community composition as the outcome, stratified by population.

|  | ELICIT |  |  | MISAME-III |  |  | MUMTA |  |  |
| --- | --- | --- | --- | --- | --- | --- | --- | --- | --- |
|  | *Sorensen dissimilarity\** |  |  | *Sorensen dissimilarity\** |  |  | *Sorensen dissimilarity\** |  |  |
| *Predictor* | *R*<sup>2</sup> * 100% | *pseudo-F* | *p-value* | *R*<sup>2</sup> * 100% | *pseudo-F* | *p-value* | *R*<sup>2</sup> * 100% | *pseudo-F* | *p-value* |
| Maternal age (years) | 1.436 | 1.788 | 0.006 | - | - | - | 0.812 | 1.139 | 0.272 |
| Parity (cont.) | - | - | - | - | - | - | 0.756 | 1.061 | 0.365 |
| Maternal BMI | - | - | - | - | - | - | 0.859 | 1.205 | 0.202 |
| Delivery location | 1.287 | 1.602 | 0.026 | - | - | - | - | - | - |
| Improved water source | 0.956 | 1.190 | 0.194 | 0.840 | 1.598 | 0.038 | - | - | - |
| Delivery season (pre vs post-harvest) | 1.302 | 1.622 | 0.035 | - | - | - | - | - | - |
| Delivery season (rainy vs non-rainy) | - | - | - | 1.302 | 2.474 | 0.001 | 1.407 | 1.973 | 0.004 |
|  | *Bray-Curtis dissimilarity* |  |  | *Bray-Curtis dissimilarity* |  |  | *Bray-Curtis dissimilarity* |  |  |
| Maternal age (years) | 1.312 | 1.641 | 0.077 | - | - | - | 0.481 | 0.698 | 0.680 |
| Parity (cont.) | - | - | - | - | - | - | 0.825 | 1.196 | 0.292 |
| Maternal BMI | - | - | - | - | - | - | 0.778 | 1.129 | 0.304 |
| Delivery location | 1.556 | 1.945 | 0.027 | - | - | - | - | - | - |
| Improved water source | 1.173 | 1.466 | 0.120 | 1.168 | 2.215 | 0.009 | - | - | - |
| Delivery season (pre vs post-harvest) | 1.215 | 1.519 | 0.104 | - | - | - | - | - | - |
| Delivery season (rainy vs non-rainy) | - | - | - | 0.717 | 1.359 | 0.161 | 4.335 | 6.288 | <0.001 |

\*Sorensen dissimilarity is an unweighted version of Bray-Curtis dissimilarity (dissimilarity based on ASV presence/absence)

Machine learning models predicted infant birth season using HM bacterial abundances with high accuracy in all three populations (AUROC > 0.70; **Figure 4a**), though the most predictive features (ASVs) varied across models (**Figure 4b-d; Table S5**). Model performance improved when birth season in ELICIT was classified as pre or post-harvest (AUROC = 0.75) rather than rainy or non-rainy (AUROC = 0.55; **Figure 4a**). On a global level, *Corynebacterium* was the strongest predictor of birth season in both MISAME-III and MUMTA (**Figure 4b-c**), while *Corynebacterium* and *Veillonella* were equally predictive of birth season in ELICIT (**Figure S3a**). Irrespective of genera abundances however, ASVs belonging to certain genera—for instance *Streptococcus* in MISAME-III and MUMTA, and *Acinetobacter, Veillonella* and *Dietzia* in ELICIT— displayed notable variation in the directionality of their contribution to the prediction of birth season (i.e. positive, negative, or minimal (values near 0)). This was particularly notable for *Corynebacterium*, which had the highest number of ASVs across the populations (25, 19, and 22 in ELICIT, MISAME-III, and MUMTA), including a number of predictive ASVs that displayed variable relationships with birth season (**Figure 4b-c**, **Figure S3a**). For example, in MISAME-III, there were three *Corynebacterium* ASVs among the top 15 most important features; *Corynebacterium* 258 abundance was depleted while *Corynebacterium 85* and *Corynebacteirum amycolatum 1* were slightly enriched in the rainy season (**Figure 4b**). In MUMTA, the two most predictive *Corynebacterium* ASVs were enriched in the rainy season (**Figure 4c**). *Corynebacterium* also had elevated richness in the rainy season (mean, 8.0 ASVs) compared to the non-rainy season (6.4, p = 0.003) in the MUMTA study, though the inverse was true for MISAME-III (5.9 rainy, 7.3 non-rainy season, p = 0.019) (**Figure 4d**). Richness did not differ by infant birth season in MUMTA or MISAME-III for any other genera assessed.

**Figure 4.**
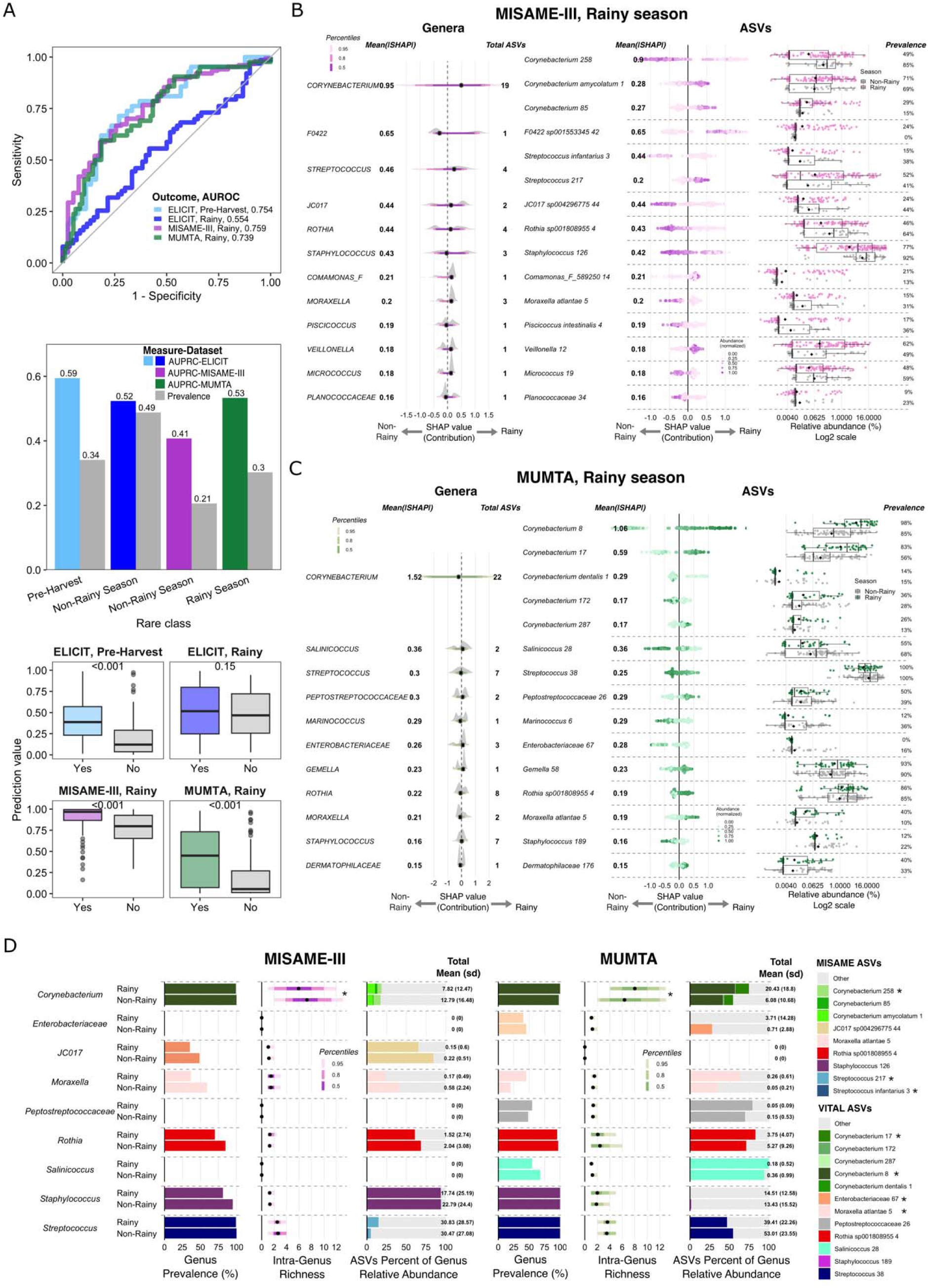
Infant birth season was consistently predicted from human milk microbiota across geographic settings. (a) Using xgboost models, human milk microbiota predicted the season of infant birth in all studies, though this was dependent on the classification of seasons. Accuracy of the prediction of the rainy season in all sites, and for the prediction of the pre-harvest season in ELICIT, was evaluated based on: (top) AUROC values for each model; (middle) AUPRC values at least greater than the prevalence of the rare group (annotated on x-axis) in each study, and (bottom) whether the estimated prediction probability was significantly greater in the true case group (pre-harvest and rainy season) compared to the control (post-harvest and non-rainy season) in each study (p-values from Wilcoxon rank-sum tests). Also see Table 4. (b-c) Feature group (genus) and individual feature (ASV) importance for the top 15 most predictive ASVs (based on the mean of absolute SHAP values) are shown for the models accurately predicting rainy season in (b) MISAME-III and (c) MUMTA. ASV relative abundances are also shown within the true rainy (colored points) and non-rainy (gray points) seasons with the median (line) and mean (black diamond) shown on boxplots (right). See Figure S3a for feature importance in the model accurately predicting pre-harvest in ELICIT. (d) Intra-genus richness and composition between infant birth in the rainy and non-rainy season in MISAME-III and MUMTA. Left to right per study: total genus prevalence (left) and within samples that contain the genera, intra-genus richness (the total number of unique ASVs from that genus within a sample) (middle) and the percent of total genus relative abundance contributed by the top 15 ranked ASVs (right). *p<0.05, Wilcoxon rank sum test. Displaying only those genera with >1 unique ASV, of those represented in the top 15 features for a given study (e.g. Unclassified Enterobacteriaceae is among the top 15 features in MUMTA, but not among the top 15 in MISAME-III, so it is not displayed in MISAME-III). See Figure S3b for this follow-up analysis on the most important features predicting pre-harvest in ELICIT.

### The HM microbiome predicted infant growth

Machine learning models predicted infant growth outcomes with variable accuracy across the three studies (**Figure 5a; Figure S4a; Table S5**). Infant stunting at three months of age was predicted with relatively high accuracy in ELICIT and MUMTA (ELICIT AUROC = 0.69, AUPRC = 0.44, p = 0.003; MUMTA AUROC = 0.72, AUPRC = 0.36, p = 0.001), where the prevalence of stunting was 19% and 20%, respectively. *Corynebacterium* contributed to the prediction of infant stunting in ELICIT and MUMTA, along with *Rothia, Veillonella* and *Acinetobacter* (**Figure 5b-5c**). *Corynebacterium* displayed similar patterns compared to those detected in the predictive models of infant birth season, with varying directionality in its contribution to the prediction of stunting among samples, as well as differences between specific ASVs within the genus.

**Figure 5.**
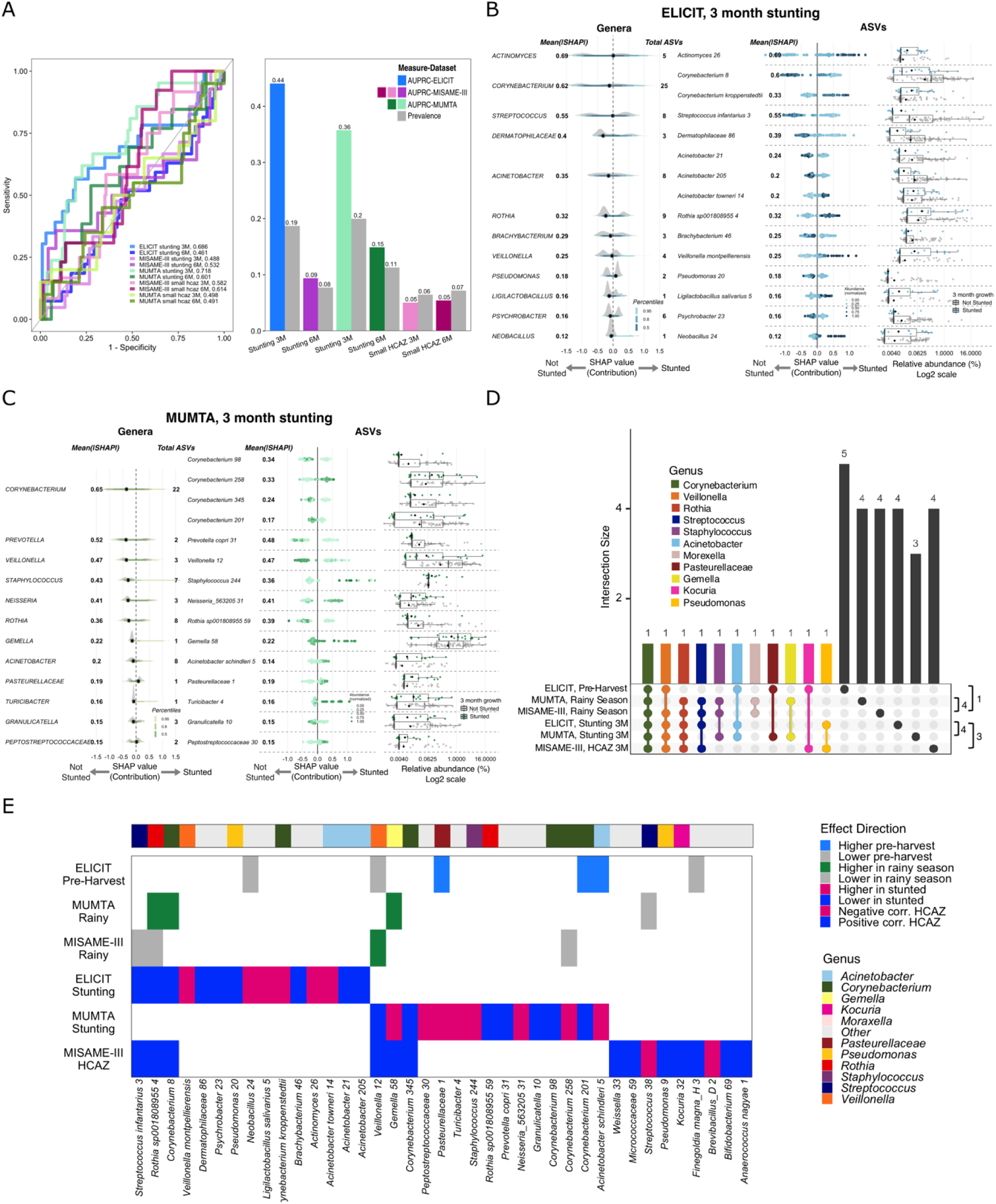
Contrast in the milk microbiota most important to the prediction of infant birth season and growth outcomes across populations. Variation in the prediction of Infant growth outcomes from human milk microbiota across geographic settings. (a) Using xgboost models, human milk microbiota predicted infant stunting at three months with weak-to-moderate accuracy depending on study and growth outcome, based on the following performance metrics: AUROC values for each model (left); AUPRC values at least greater than the prevalence of the rare group (poor growth outcome) in each study (right), and whether the estimated prediction probability was significantly greater in the true case group compared to the control in each study, based on Wilcoxon rank-sum tests (see Figure S4A). Also see Table 4. (b & c) Feature group (genus) and individual feature (ASV) importance for the top 15 most predictive ASVs (based on the mean of absolute SHAP values) are shown for the models accurately predicting stunting at 3 months in ELICIT (b) and MUMTA (c). Also see Figure S5c for feature importance of HCAZ at 3 months in MISAME-III, and Figure S4b for additional follow-up analyses of the most important ASVs for the prediction of stunting at 3 months. (d) Overlap in the 10 genera most predictive of infant birth season and growth outcomes. The dendrogram annotation to the right shows the total overlap for similar outcomes (e.g. for the prediction of rainy season birth, 4 of the top 10 genera overlapped between MUMTA and MISAME-III. Only the 6 outcomes accurately predicted from human milk microbiota are shown. Feature group importance is based on the mean of absolute SHAP values. (e) The relationship between key ASVs and infant birth season and growth outcomes (outcomes shown on the y-axis, cell is shaded if ASV is in the top 15 for that model based on the mean of absolute SHAP values). The color shade indicates whether the ASV was in a higher or lower abundance in samples associated with each outcome. The plot includes only those features most important to infant growth predictions.

*Actinomyces* was uniquely important to the prediction of infant stunting in the ELICIT population, where it had the highest importance ranking, whereas *Prevotella* was uniquely important to the prediction of stunting in MUMTA (**Figure 5b-5c**). We additionally aimed to highlight within-genus variation, where potentially rarer ASVs differ from the typical genus pattern. Among genera with > 1 ASV, we identified several instances where ASVs most predictive of infant stunting did not make up a large proportion of total genus relative abundance and also displayed differences in dominance patterns (**Figure S4b**). In ELICIT, *Actinomyces 26* and *Corynebacterium kroppenstedtii* were enriched in HM from mothers of stunted infants (22.85% and 6.0%, respectively) compared to those with non-stunted growth (9.74%, p=0.012 and 3.0%, p = 0.026), while *Acinetobacter 205* and *Corynebacterium* 8 were depleted (8.7% vs. 15.1%, p=0.022 and 5.3% vs. 17.9%, p=0.006). There were also important ASVs in MUMTA that made up a low proportion of genus abundance.This included two *Corynebacterium* ASVs (98 and 345) depleted in HM from mothers of stunted infants (0.04% vs. 1.5%, p = 0.047 and 0.75% vs. 4.73%, p = 0.046).

In contrast to findings in ELICIT and MUMTA, the models poorly predicted infant stunting in MISAME-III (**Figure 5a; Table S5**), where prevalence was 11%. We instead applied a similar xgboost algorithm to perform regression for continuous growth outcomes. This algorithm accurately predicted head circumference-for-age z-score (HCAZ) at three months in MISAME-III (**Figure S5a**, Spearman rho [95% CI] = 0.17 [0.06-0.28], p<0.001), though no outcomes were accurately predicted in ELICIT or MUMTA. Correlations between HM ASV abundances and continuous infant growth metrics varied by population and infant birth season, but in line with model results, the strongest correlations were detected for HCAZ in MISAME-III (**Figure S5b; Figure S6**). The most predictive feature in the model, *Anaerococcus nagyae.1*, was strongly positively correlated with HCAZ at both three and six months of age (**Figure S5b & 6c**), and the most significant positive correlations at 3 months persisted to 6 months (7 of 8 correlations).

These patterns displayed a signal of seasonality, as strong positive correlations to infant growth were exclusive to the milk of mothers who delivered during the rainy season in MISAME-III. Correlations were much weaker, and not statistically significant, in the milk of mothers who delivered in the non-rainy season (**Figure S5b**). Correlations in ELICIT and MUMTA were generally weaker (**Figure S6**).

### Consistency in the key contributors to the prediction of birth season and infant growth

At the level of genus, *Corynebacterium* was a key contributor to the prediction of birth season and infant growth outcomes across all populations (**Figure 5d**). *Veillonella* and *Rothia* were additional contributors to predictions of infant growth and rainy season birth. *Acinetobacter* predicted stunting at three months in MUMTA and ELICIT, as well as pre-harvest season birth in ELICIT. At the individual feature level, the model predictors were less consistent. However, we identified some HM ASVs with significant associations to infant growth across two or more populations (**Figure 5e**). For example, *Veillonella_A.12* and *Corynebacterium.345* correlated positively with infant HCAZ in MISAME-III and had a lower abundance in HM of MUMTA mothers of stunted infants. Similarly, *Streptococcus infantarius*, *Rothia spp4.*, and *Corynebacterium.8* correlated positively with infant HCAZ in MISAME-III and were lower abundance in HM from mothers of stunted infants in ELICIT. Despite overall consistency in *Corynebacterium* as an important contributor in all six models, the individual features most important to predictions frequently differed, in both ASV-level identity and directionality, across populations and predicted outcomes.

## Discussion

This study highlighted commonalities, as well as key differences, in the HM microbiome across distinct geographies, environments, and lifestyles. Our data revealed associations between the HM microbiome and infant growth, and showed that *Corynebacterium* was a global predictor of infant growth phenotypes. Differences in the specific *Corynebacterium* ASVs that predicted infant growth suggests this phylogenetically diverse genus contains species with functional overlap, thus yielding differences in the directionality of relationships between *Corynebacterium* ASVs and infant growth across the populations. We also found seasonal variation in the HM microbiome, highlighting its sensitivity to local environmental factors. These findings contribute to the growing literature on the milk microbiome of humans (Williams et al. 2017; Meehan et al. 2018; Lackey et al. 2019; Ma et al. 2024) and other primates (Muletz-Wolz et al. 2019; Bornbusch et al. 2024, 2022), and shine an evolutionary light on the HM microbiome as a biological system that embeds local environmental factors and shapes key phenotypes related to infant survival.

### HM microbiome varied by geography yet three bacterial genera were detected in all samples

All HM samples contained five “core” ASVs belonging to *Corynebacterium*, *Staphylococcus*, and *Streptococcus*, a finding in line with other studies that detected *Corynebacterium* in HM across global populations (Williams et al. 2017; Lackey et al. 2019; Meehan et al. 2018) and a high prevalence of *Streptococcus* and *Staphylococcus* in non-human primate milk (Muletz-Wolz et al. 2019). We also found uniform relative abundances of core *Streptococcus* and *Staphylococcus* ASVs across the populations. As these genera are human skin commensals, their elevated prevalence in HM may reflect seeding by microbes originating on maternal and/or infant skin, rather than a source of contamination. This is likely the result of direct contact between mother-infant dyads during breastfeeding (Meehan et al. 2018; Ma et al. 2024), as previously demonstrated by an elevated prevalence of skin commensals in the HM of exclusively breastfeeding Canadian mothers compared to those who breastfed and pumped (Fehr et al. 2020).

We also detected a clear geographic signal in the HM microbiome. ELICIT samples displayed the highest bacterial diversity and richness, including ASVs not detected in MISAME-III or MUMTA, and the lowest relative abundance of core ASVs *Corynebacterium amycolatum.1* and *Corynebacterium.8*. Across the study sites, variation in diet, hygiene practices, and contact with bacteria in the natural environment may promote differential transmission of soil and water-associated taxa (Balows et al. 2013; Sun et al. 2018; Mujumdar et al. 2023) to HM via enteromammary transmission (Rodríguez 2014) and/or retrograde inoculation (Ramsay et al. 2004; Moossavi and Azad 2020). To this point, we found that household water source explained HM microbiome variation in ELICIT and MISAME-III. While differences in sample collection protocols across the studies may contribute to the observed variation, our findings echo broader evidence that the local environment shapes microbial communities in human (Lackey et al. 2019) and non-human primate (Bornbusch et al. 2024) milk and other body sites (Rosas-Plaza et al. 2022; Blaser et al. 2013; Manus et al. 2020). Our findings suggest that local ecology contributes to HM microbiome profiles, and guides future research to sample local ecosystems to better understand how microbes move through the mother-infant-milk triad (Shenhav and Azad 2022)—an evolutionarily relevant system with lasting effects on health.

### Core genera predicted infant growth, with variation in specific ASVs

*Corynebacterium* was a key predictor of infant growth in each study. The relative abundances of two *Corynebacterium* ASVs were positively correlated to infant HCAZ in MISAME-III and depleted in association with infant stunting in ELICIT and MUMTA, indicative of a beneficial relationship. However, enrichment of two different *Corynebacterium* ASVs was associated with infant stunting in ELICIT and MUMTA. This mirrors contrasting findings in previous studies, including negative associations between HM *Corynebacterium* relative abundance and infant HCAZ in Guatemala (Ajeeb et al. 2022) and positive associations between *Corynebacterium* relative abundance in the infant oral microbiome and improved growth in the USA (Coker et al. 2022). Thus, a “healthy” HM microbiome is likely context-specific, reflecting differences in both the environmentally-sourced microbes that colonize maternal and infant bodies, as well as the health challenges experienced by mothers and infants across settings. Though not indicative of causal relationships, our results identify *Corynebacterium* as a potential candidate for interventions that target the microbiome to improve infant growth. For example, promoting ASVs associated with healthy growth could benefit infants in settings where elevated immune activity due to infection diverts energy away from growth, thus contributing to growth faltering (Urlacher et al. 2018; Garcia et al. 2020).

We also detected ASV-level variation in other predictive genera. The relative abundance of core ASV *Streptococcus.38* correlated negatively to infant HCAZ in MISAME-III, while phylogenetically related *Streptococcus infantarius.3* was positively correlated to the same outcome. HM *Streptococcus* was previously linked to higher HCAZ in Guatemalan infants (Ajeeb et al. 2022). Further, *Veillonella.12* was associated with positive growth outcomes in MISAME-III and MUMTA, while enriched *Veillonella montpellierensis* was associated with infant stunting in ELICIT. Previously, *Veillonella* in the pediatric oral microbiome was linked to stunting in Indonesia (Theodorea et al. 2022). Intra-genus variation in relationships to infant growth in the current study may reflect meaningful differences in host-ASV relationships, though more work is needed to understand when taxonomic diversity begets functional diversity that impacts maternal and infant health.

### Core genera were associated with infant birth season

Replicating findings in Australia (Ma et al. 2024), we detected seasonal variation in the HM microbiome, with *Corynebacterium* as a strong predictor of infant birth season in each population. Multiple *Corynebacterium* ASVs displayed elevated abundances in samples from MUMTA women who gave birth in the rainy season (compared to non-rainy season) and ELICIT women who gave birth before the harvest (compared to after). In contrast, the core ASVs *Streptococcus.38* and *Staphylococcus.126* were depleted in the HM of mothers who delivered in the rainy season in MUMTA and MISAME-III (compared to non-rainy season). This suggests that the abundances of prevalent HM bacteria are sensitive to variable temperature, humidity (Jensen et al. 2023), and sun exposure (Harel et al. 2023), along with seasonal variation in maternal diets (Hanley-Cook et al. 2022) and hygiene practices. These seasonal differences in the HM microbiome may have downstream impacts on infant microbial colonization, in line with a study in Botswana where *Corynebacterium* enrichment in the nasopharyngeal microbiome of HM-fed infants also displayed seasonal variation (Kelly et al. 2022). It is noteworthy that the strong signal of seasonality persisted across the IMiC populations, despite distinct climates and weather patterns. Akin to the gut and skin microbiomes (Callewaert et al. 2020; Davenport et al 2014), sensitivity to the local environment may be a global phenomenon of the HM microbiome, supporting its evolutionary importance as a system that directly impacts infant health by embedding factors of the local environment.

### Strengths and limitations

Our study is the first to combine a large dataset of HM samples collected from international populations with detailed environmental data and infant anthropometry. This rich dataset enabled the use of machine learning techniques that identify seasonal variation in the HM microbiome and taxa most predictive of early life growth. Though we took multiple steps to standardize laboratory and computational analysis, account for potential contamination, and reduce noise due to infant age (Lönnerdal et al. 2017), it is possible that some of the inter-population variation that we detected is amplified by differences in sample collection procedures across the studies. Further, we were unable to incorporate data on maternal diets or the immune phenotypes of mother-infant dyads, factors likely to shape the bacterial composition of HM.

While *Corynebacterium* was a global predictor of infant growth across the IMiC studies, we detected variation at the level of ASV. This suggests that phylogenetically related ASVs may encode similar functional pathways that impact infant growth across distinct geographies.

Previous work that used the same predictive modeling technique found functional metagenomic features of infant stool bacteria better predicted linear growth compared to taxonomic features alone (Robertson et al. 2023). Methodological advancements are needed to apply metagenomic sequencing to low-biomass HM samples.

## Conclusion

We identified for the first time HM bacteria with strong associations to infant growth across three geographically and culturally distinct populations where undernutrition and growth faltering is of concern. We also corroborate previous findings of seasonal variation in the HM microbiome.

Moving forward, a systems biology approach will help untangle the ecological interactions between HM bacteria and nutrients (LeBlanc et al. 2013), as well as bacteria in HM and the infant gut, that likely influence infant growth in the IMiC cohorts. Since the microbial genome can extend human phenotypic plasticity (Dunn et al. 2020), we advocate for future work that identifies connections between HM bacterial gene pathways and infant physiology. This will expand on our findings of the HM microbiome as an evolutionary relevant system that is environmentally sensitive, shared across generations, and associated with infant growth phenotypes.

## Methods

### Cohort demographics and study details

We analyzed 451 HM samples collected from 451 mothers living in rural Tanzania, rural Burkina Faso, and peri-urban Pakistan, between one and two months postpartum. While the larger IMiC study includes HM samples and anthropometric data collected at multiple timepoints per population, as well as samples collected from mothers living in Canada, this manuscript focuses on HM samples that were collected at comparable timepoints across the three low-to-middle income countries included in IMiC (**Table 1**). Participant eligibility and enrollment, as well as additional details for each of the three studies analyzed in the current manuscript, can be found on ClinicalTrials.gov and in the protocol paper published by each study team (ELICIT: NCT03268902, (DeBoer et al. 2018); MISAME-III: NCT03533712, (Vanslambrouck et al. 2021); MUMTA: NCT03564652, (Muhammad et al. 2020)). Country-level sociodemographic and nutrition indicators and inclusion criteria for each IMiC study, as well as participant data cleaning and harmonization, are summarized in the IMiC protocol paper (Fehr et al. 2025). All participants provided informed consent and individual studies were approved by local research ethics boards at the respective lead institutions. The IMiC Consortium is funded by the Bill & Melinda Gates Foundation, is registered at ClinicalTrials.gov (NCT05119166) and was approved by the Human Research Ethics Board at the University of Manitoba.

### Infant anthropometric and birth season derivations

Questionnaire and clinical data harmonization across the studies is described in the IMiC protocol paper (Fehr et al. 2025). Briefly, infant weight, length and head circumference were measured at 3 and 6 months of age across all studies. Anthropometric z-scores were derived using the WHO child growth standards (Bloem 2007), resulting in length-for-age (LAZ), weight-for-age (WAZ), and head circumference-for-age (HCAZ). Infant stunting was defined as LAZ<-2 standard deviations below the mean (Schwarz et al. 2008). For the derivation of the birth season variable, we used local definitions of seasons within each study. In MUMTA, the monsoon season, spanning from July through September in Pakistan, was categorized as ‘rainy season,’ whereas the rainy season in Burkina Faso, from May through September, was used as the ‘rainy season’ for MISAME-III. In Tanzania, the seasons were categorized based on the local crop harvest, with April through October as ‘post-harvest’ and November through March as ‘pre-harvest’ in ELICIT. For direct comparison with MUMTA and MISAME-III, we created a second season variable for ELICIT based on average rainfall throughout the year, categorizing January through April as the ‘rainy season’.

### HM sample collection

The protocols for HM sample collection varied across IMiC studies. In the MISAME-III study, mothers provided a fully expressed HM sample from a cleaned breast with a pump. In the ELICIT study, 8 mL of milk was hand-expressed mid-feed, and in the MUMTA study, at least 10 mL of milk was hand-expressed immediately prior to breastfeeding, rather than full expression. In all three studies, the breast used for an infant’s last feeding was not used for sample collection. HM was collected in sterile containers and transported on ice to a central laboratory where it was stored at –80 C, following homogenization and splitting into 3 to 4 aliquots (Fehr et al. 2025).

### DNA extraction and microbiome sequencing

DNA extraction and microbiome sequencing was performed at the Alkek Center for Metagenomics and Microbiome Research (CMMR) at the Baylor College of Medicine. HM samples (1 to 1.5 mL) were spun at 10,000 xg for 5 minutes, and fat and supernatant was removed. The remaining milk pellet was resuspended in 800 uL of CD1 buffer from the DNeasy 96 PowerSoil Pro HT extraction kit, which was then used to complete DNA extraction following the manufacturer’s protocol on the Qiagen QIAcube HT automated extraction platform. The methods for 16S rRNA gene sequencing were adapted from the NIH-Human Microbiome Project (Human Microbiome Project Consortium 2012) and the Earth Microbiome Project (Thompson et al. 2017). Briefly, the V4 hypervariable region of the 16S rDNA region was amplified using 515F (GTGCCAGCMGCCGCGGTAA) and 806R (GGACTACHVGGGTWTCTAAT) Illumina adapted primers (single-index barcodes incorporated into the reverse primer). Paired-end amplicon sequencing was performed on the Illumina MiSeq platform. Sequences were demultiplexed based on the single-index barcodes using the Illumina ‘bcl2fastq’ software. Quality filtering of demultiplexed read pairs using bbduk.sh (BBMap, version 38.82) removed the Illumina adapters, PhiX reads, and reads with a Phred quality score <15 and length <100 bp after trimming. DNA extraction and sequencing used negative controls (DNA free water), and technical replicates (a pooled HM sample) were distributed across analysis plates and sequencing runs to assess contamination and technical variation (**Figure S1a**).

### Data pre-processing

Data were processed in the THRiVE Discovery Lab at the University of Manitoba, Canada, using methods adapted from previous studies of the HM microbiome (Fehr et al. 2020); (Moossavi et al. 2021), with some modifications (**Figure S1b**). Overlapping paired-end reads were processed with the *DADA2* v1.26.0 pipeline using the open-source software QIIME2 v2023.2 (https://qiime2.org) through the Digital Research Alliance of Canada (Callahan et al. 2016; Bolyen et al. 2019). Across the 2,184 HM samples collected from all IMiC populations and timepoints, a total of 49,033 amplicon sequence variants (ASVs) passed *DADA2* denoising.

Taxonomy was assigned using a Naive Bayes classifier trained on the GreenGenes2 v2022.10 reference database (McDonald et al. 2024). We compared these taxonomic assignments to those generated with the SILVA v138 database (Quast et al. 2013). While GreenGenes2 yielded a higher proportion of ASVs assigned at the species level in our dataset (35% compared to 10% of ASVs using SILVA), there were 2,230 ASVs that were unassigned at the phylum level with GreenGenes2, yet were assigned using SILVA. Therefore, though the majority of the original 49,033 ASVs were assigned using GreenGenes2, a total of 2,230 ASVs were assigned using the SILVA database. Since all ASVs assigned to *Cyanobacteria* in GreenGenes2 were identified as chloroplast-associated DNA using SILVA, the reads assigned to *Cyanobacteria* were also removed.

After removal of ASVs identified as contaminants and further data cleaning using the *SCRuB* (Austin et al. 2023), *decontam* (Davis et al. 2018), and *phyloseq* (McMurdie and Holmes 2013) packages in R (R Core Team 2024), samples with fewer than 8,000 reads were removed based on rarefaction curves. This threshold has been used in previous studies of the HM microbiome (Fehr et al. 2020; Fang et al. 2024). Spurious ASVs with mean relative abundance < 0.0001% were also removed, leaving 7,234 ASVs that made up the majority of total reads after contaminant removal (median [IQR], 99.7% [99.3-99.9] of total reads). For alpha-diversity only, samples were rarefied to the minimum sample depth (8,000 reads) to ensure diversity and richness metrics were comparable between samples.

Non-rarefied data were used for other analyses (e.g. of bacterial community composition and ASV differential abundance), with additional filtering to remove ASVs present in fewer than 10% of samples on the data subset the statistical test was performed on (unless specified otherwise in the statistical analysis section). For instance, for comparisons across sites, the filter was applied to the whole dataset, and for within-site tests, the filter was applied to each site separately. This minimized issues with data sparsity after subsetting the data. In cases where data were centered-log ratio (CLR) transformed, the transformation was done using the *microbiome* package (Lahti, Shetty, and Salojarvi 2017), after the data subsetting and feature filtering, and after data was zero imputed using a Bayesian-Multiplicative replacement method from the *zCompositions* package (McMurdie and Holmes 2013).

### Statistical analyses

All statistical analysis and data visualizations were generated using R software v4.4.1 (R Core Team 2024).

### Comparisons of HM microbiome profiles across populations (Q1)

We used permutational multivariate analysis of variance (PERMANOVA) of Bray-Curtis dissimilarity (using relativized data) and Sorensen dissimilarity (using presence/absence data) to test for differences in bacterial community composition across populations. Models were constructed in the *vegan* package (Oksanen, Simpson, and Blanchet 2024), and data were visualized using non-metric multidimensional scaling (NMDS) in the *ggplot2* package (Wickham 2009).

The core microbiome of each study population (ASVs present in ≥60% of samples per-site) was visualized using UpSet plots (*UpSetR* package (Gehlenborg 2019)). Data were relativized and all ASVs present in ≥60% of samples in at least one site were visualized across sites in stacked bar plots (*ggplot2* package, (Wickham 2009)). Wilcoxon rank-sum tests detected differences in Shannon diversity and bacterial richness (observed ASVs) across study populations. Shannon diversity and bacterial richness were visualized with boxplots using *ggplot2* (Wickham 2009).

Differential ASV abundances were tested in pairwise comparisons between populations using *ANCOM-BC2* (Lin, Eggesbø, and Peddada 2022), which applied a pseudo-count to zero values followed by a log transformation to the count data. To avoid structural zeros, only ASVs present in at least 10% of samples within both sites being compared were tested (for comparisons of MISAME-III-ELICIT, MUMTA-ELICIT and MISAME-III-MUMTA, 80, 134, and 78 ASVs were tested, respectively). P-values were adjusted for multiple comparisons using the Benjamini-Hochberg (BH) procedure. The total number of significant differences (p(BH)<0.05) identified among comparisons were visualized in UpSet plots (*UpSetR* package (Gehlenborg 2019)), and we used the *ggplot2* package to display log-fold differences for ASVs that were the most differentially abundant (p(BH)<0.0001) in at least one of the between-site comparisons (Wickham 2009).

We also calculated intra-genus richness as the number of unique ASVs from a given genus within a sample. Samples that lacked ASVs from a given genus were excluded, such that genus richness values were not dependent on prevalence of the genus (i.e. instead of coding richness as 0 whenever the genus was completely absent, the sample was instead excluded), and genera including a single ASV were excluded. Genus richness was visualized with interval plots using *ggdist* and *ggplot2* packages (Kay 2024; Wickham 2009), and compared between sites using Wilcoxon rank-sum tests, with p-values adjusted using the BH procedure.

### Assessment of factors associated with the HM microbiome within populations (Q2)

We performed a redundancy analysis (RDA) using the *vegan* package to identify the strongest predictors of overall HM bacterial abundance profiles in each population (Oksanen, Simpson, and Blanchet 2024), where the outcome was the overall ASV abundance profile (CLR-transformed abundances). Within each study, a univariable RDA was first performed for each of the predictor variables of interest, followed by a multivariable RDA that included the significant predictors identified in the univariable models. Partial RDA (pRDA) was used to determine the variation explained by infant birth season after the removal of the effects of covariates in multivariable models. The complete list of univariable predictors for the models included: infant sex; gestational age (days); infant age (days); maternal age (years); number of people living in the household; parity; maternal BMI; improved water source (i.e. tap water); improved flooring (i.e. non-clay); infant birth location; infant birth season; and any infant diarrhea, cough, or vomit reported by the mother between birth and the study visit. Not every variable was available in each of the studies (**Table 1**). Within each study, infant birth season was categorized as rainy vs non-rainy season. Based on the importance of agricultural practices in Tanzania, the infant delivery season variable was additionally categorized as ‘pre-harvest vs post-harvest’ in the ELICIT study.

To test if the significant predictors of variation in bacterial abundance profiles in the RDA were also predictors of variation in bacterial community composition based on beta diversity metrics, we used PERMANOVA of Bray-Curtis dissimilarity and Sorensen dissimilarity (Oksanen, Simpson, and Blanchet 2024). The predictors selected for this multivariate analysis were those identified as significant in the univariate RDAs within each study.

### Investigating infant birth season as a key predictor of HM microbiome profiles (Q2)

We then focused on infant birth season, the factor that was consistently associated with HM microbiota composition and abundance profiles within each population based on the analyses described above. For alpha diversity and differential abundance analyses of birth season within populations, we used the same approaches as between-population comparisons. Briefly, Wilcoxon rank-sum tests were used to test for differences in Shannon diversity and bacterial richness between seasons, and for ASVs present in at least 10% of samples within each population, *ANCOM-BC2* was used to test for differences in abundance of ASVs between seasons (Lin, Eggesbø, and Peddada 2022).

### Prediction of infant birth season (Q2) and growth outcomes (Q3) using HM microbiota profiles

To predict infant birth season (Q2) and growth outcomes (Q3) using CLR-transformed ASV abundances, we used the *xgboost* R package to construct Extreme Gradient Boosting (XGBoost) machine learning models (Chen et al. 2024; Chen and Guestrin 2016). XGBoost implements gradient boosted decision trees, and was used to efficiently capture complex nonlinear relationships between microbiota and outcomes. Models were validated using 10 repetitions of a 10-fold cross-validation strategy (Stone 1976). Splits were stratified by the outcome variable using the ‘creatFolds’ function of the *caret* package (Kuhn 2008). For continuous infant growth outcomes, the performance of regressions was evaluated by assessing the Spearman ρ and a p-value < 0.05 to assess a positive monotonic association between the model’s predicted outcome from test data, and the true outcome values. The Spearman correlation metric was averaged across repetitions, and a 95% confidence interval was calculated. Similarly, performance of classification models was evaluated using model prediction scores calculated from the test data of each fold, aggregated, and averaged across repeats to avoid inflation and reduce error in performance estimates. Performance criteria for the classification models included: (i) an area under the receiver operating characteristic curve (AUROC) value greater than 0.5; (ii) an area under the precision-recall curve (AUPRC) value greater than the prevalence of samples in the “rare class” (i.e. binary category with the smaller sample size); and (iii) p-value < 0.05, generated from a Wilcoxon rank-sum test assessing whether the prediction probability was greater for the true case group compared to the control. A model needed to pass all three of these criteria in order for performance to be considered sufficient for feature attribution analysis. The intersection of these criteria was used to increase robustness of model evaluation. We emphasized the AUPRC value as it provides a more accurate estimate of model performance when using imbalanced datasets (i.e. disparity between the number of samples in the case and control groups), as the calculation gives more weight to the minority, or rare, class (Shu et al. 2024).

The models that passed performance metric thresholds were considered accurate, and further evaluated for interpretation on a per-model basis. Specifically, the importance of individual ASVs to the model prediction was assessed using Shapley Additive Explanation (SHAP) values (Lundberg, Scott M., and Su-In Lee 2017), which were calculated using the ‘predict’ function of the *xgboost* package. SHAP values were derived by comparing the model’s predicted outcome with and without the given feature, for approximately all possible combinations of feature subsets. SHAP values represent the marginal relative contribution of a feature (i.e. ASV) to the predicted outcome of a sample, accounting for both the individual feature and its possible coalitions with other features. Global feature importance was assessed by averaging absolute SHAP values across samples. Based on this metric, the top 15 most important ASVs to the model’s prediction were visualized. Beeswarm plots illustrated an ASV’s normalized abundance (model input value) in a sample as point color shading, and the ASV’s contribution to the predicted outcome of the sample on the x-axis position.

Given that SHAP values, plus a baseline value (model prediction without taking any features into account), sum to the total marginal prediction score of a given sample, SHAP values can be summed across features per-sample while maintaining their utility. Therefore, we summed SHAP values across ASVs per-sample, grouping them at the taxonomic level of genus. This provided an additional summary of the relative contributions of bacterial genera to the predicted outcome for each model. These derived SHAP values were treated similarly to feature-level values as described above. For instance, global genus (feature group) importance was assessed averaging absolute SHAP values across samples. Any genus assigned to the top 15 most important ASVs were visualized alongside these ASVs (beeswarm plots described above). The distribution of genus-level SHAP values across samples was visualized using ridgeplots (Wickham 2009). Ridgeplots are density curves, overlayed with an interval plot showing the range in which 50, 80 and 95 percent of samples fall within (*ggdist* package, (Kay 2024)). The overall genus importance was annotated on plots, along with the total number of ASVs within each genus, since this is important reference information when contrasting with ASV-level plots. For instance, for genera containing only a single ASV, genus-level plots were simply a different representation of the same ASV-level SHAP value data. All model performance metric, feature attribution, and relative abundance visualizations were generated using *ggplot2* (Wickham 2009).

### Further characterization and interpretation of the relationships between season of delivery and the HM microbiome, and the HM microbiome and infant growth outcomes

For each classification model that passed performance metrics, we performed two follow-up analyses to gain information about the more ‘diverse’ genera represented among the top 15 most predictive ASVs, as well as the ASVs of most importance to model predictions within each of the top genera. Among the top 15 most predictive genera that contained more than one ASV (on a per-model basis), we first compared intra-genus richness between outcome groups using Wilcoxon rank-sum tests. The intra-genus richness calculation is described above. In summary, ASV richness within each genus was visualized in interval plots using the *ggdist* and *ggplot2* packages (Kay 2024; Wickham 2009). We also compared the percent of total genus relative abundance that was accounted for by each of the ASVs between outcome groups using Wilcoxon rank-sum tests. The mean percent of total genus relative abundance across samples was also shown in stacked bar plots, relative to the total genus abundance (with ASVs in the genus that were not among the top 15 most important to the outcome shaded in grey). In both follow-up analyses, only samples containing any ASV (and any reads) from the genus were included in the statistical test and visualized. Therefore, for reference to sample sizes used, total genus prevalence is also shown alongside visualizations for these analyses. Of note, the percentage of total genus relative abundance that the top 15 most important ASVs account for was used to assess how the most important ASVs ranked in terms of their abundances, relative to the other ASVs within the same genus.

We additionally performed Spearman correlations between CLR-transformed ASV abundances and four continuous infant growth outcomes (LAZ, WAZ, WHZ, and HCAZ) at three and six months of age in each population. Heatmaps of correlations were generated using *ggplot2* (Wickham 2009) and *ggnewscale* packages (Campitelli 2025), with samples stratified by infant delivery season. For ELICIT and MISAME-III, the heatmaps included ASVs with at least one correlation with a magnitude ≥0.20 or a Benjamini-Hochberg adjusted p-value <0.3. The heatmaps of correlations in MUMTA included ASVs with at least one correlation with a magnitude ≥0.20 or an unadjusted p-value < 0.01 (only 4 ASVs were retained using the filter of adjusted p-value <0.3). The top row of each heatmap included cells that display the prevalence of each ASV, with “low” indicating ASVs that were excluded from analysis due to prevalence <0.1.

Finally, we summarized feature attribution results across all ML models that passed performance metric thresholds. We used an UpSet plot to visualize the level of overlap in the identities of the top 10 most important genera to the prediction across populations and outcomes, as well as a heatmap displaying the level of overlap in the top 15 most important individual ASVs for the prediction of each outcome. The UpSet plot was generated using the *UpSetR* package (Gehlenborg 2019), while the heatmap was generated using the *ggplot2* package (Wickham 2009). In both cases, feature importance was determined as described above, using the absolute SHAP values averaged across samples. For the ASV-level heatmap, directionality was based on whether the ASV had higher or lower abundance in each outcome group. For binary outcomes, this was determined by the difference in the median CLR transformed values between the outcome groups, while Spearman correlation coefficients were used for the continuous outcome.

## Data and code availability

Microbiome sequence data are available at NCBI BioProject: PRJNA1292286.

## Data Availability

Microbiome sequence data are available at NCBI BioProject: PRJNA1292286. All data produced in the present study are available upon reasonable request to the authors.

## Acknowledgements

The authors thank the study participants and project staff in Tanzania, Burkina Faso, and Pakistan, all of whom were central to this project. The authors are also grateful for the assistance of personnel at the THRiVE Discovery Lab at the University of Manitoba and the Alkek Center for Metagenomics and Microbiome Research (CMMR) at the Baylor College of Medicine for their support of laboratory analyses and sequencing, respectively. This project also benefited from resources provided by the Digital Research Alliance of Canada (alliancecan.ca), including the Compute Canada advanced research computing system.

---

**Figure S1.**
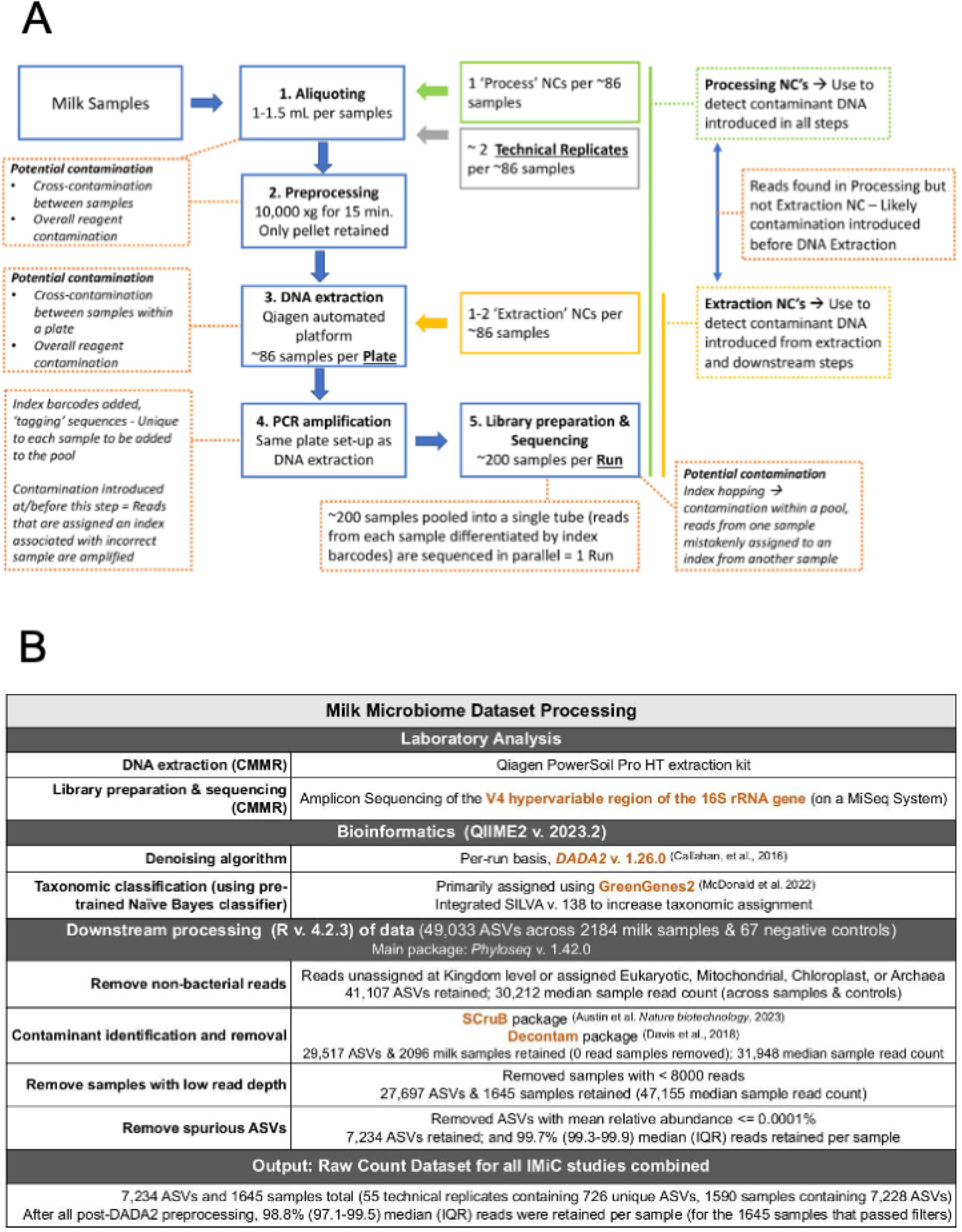
(a) Flow chart describing the plates, runs, and technical controls used in microbiome analysis, as well as potential routes of contamination that were adjusted for using the *SCruB* and *decontam* packages. (b) Milk microbiome dataset preprocessing to generate the untransformed count dataset. ASV, Amplicon Sequence Variant (1 taxonomic unit). Note that removing samples with fewer than 8,000 reads allowed the retention of 78% of all samples (though this varies slightly across the studies). We do not expect human milk samples with read counts below 8,000 reads to yield accurate microbiome richness (number of ASVs in a sample) and diversity estimates.

**Figure S2.**
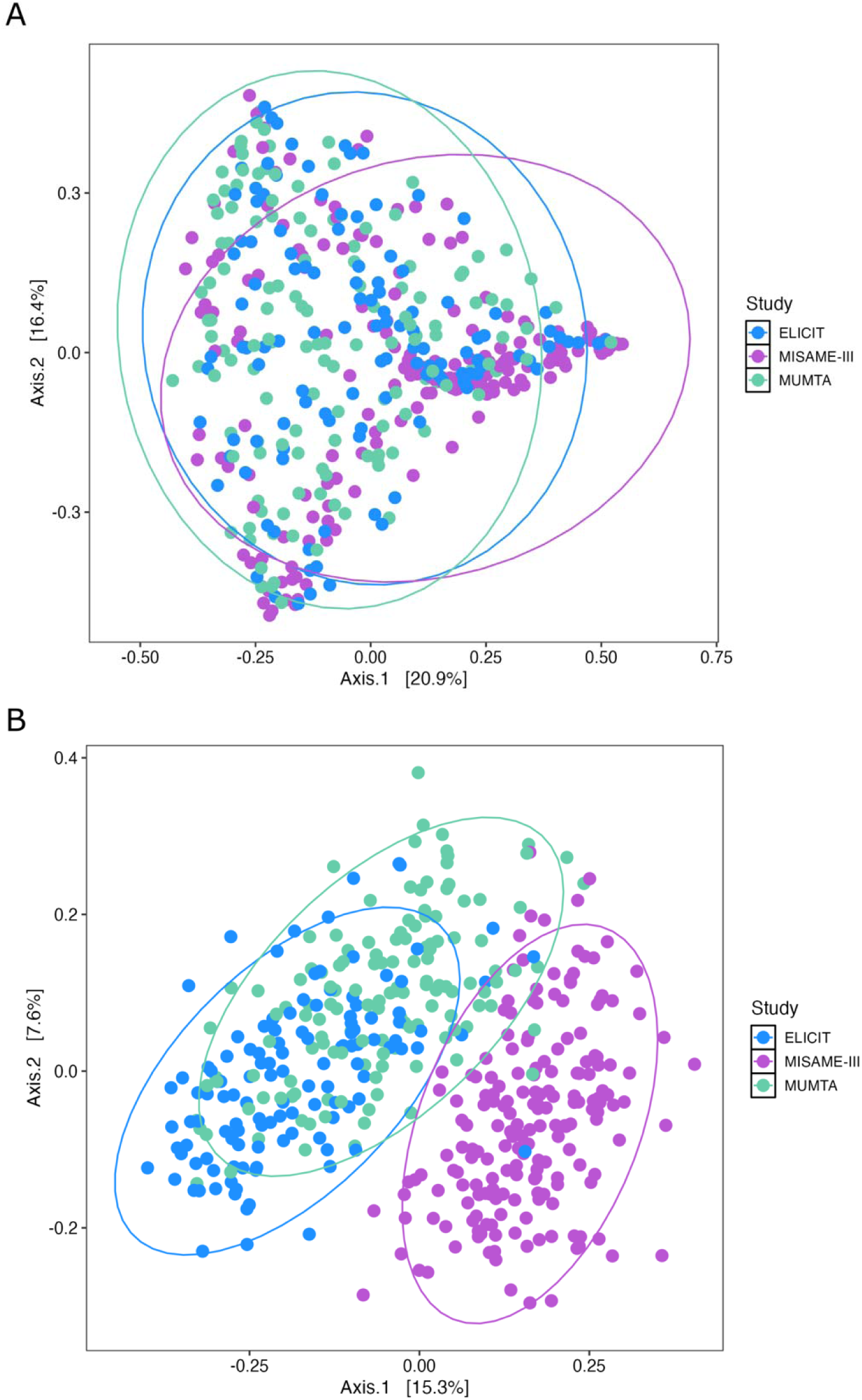
NMDS plot of (a) Bray-Curtis dissimilarity and (b) Sorensen dissimilarity (unweighted version of Bray-Curtis dissimilarity). Ellipses reflect 95% confidence around centroids. See Table 2 for accompanying statistics.

**Figure S3.**
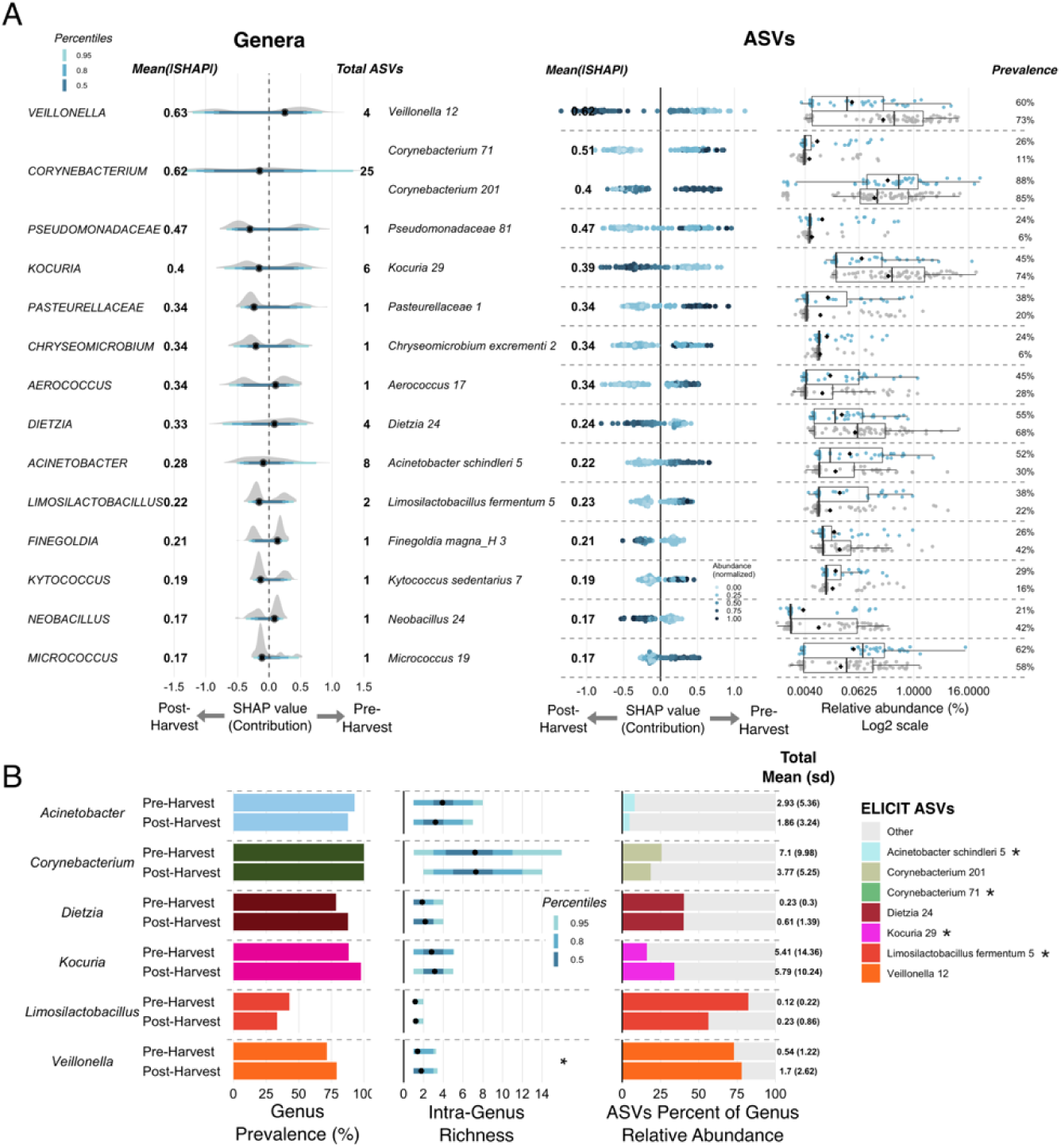
Human milk microbiota most predictive of the pre-harvest infant birth season in ELICIT. (a) Feature group (genus) and individual feature (ASV) importance for the top 15 most predictive ASVs (based on the mean of absolute SHAP values) are shown for the three models that passed all performance criteria, for the prediction of pre-vs post-harvest birth in ELICIT, ASV relative abundances are also shown within the true samples associated with the pre-harvest (colored points) and post-harvest (gray points) infant birth seasons, with median (line) and mean (black diamond) shown on boxplots (right). (b) Total genus prevalence (left) and within samples that contain the genera, intra-genus richness (the total number of unique ASVs from that genus within a sample) (middle), and the percent of total genus relative abundance contributed by the top 15 ranked ASVs (right). *p<0.05, tested using Wilcoxon rank sum test, displaying genera with ≥1 unique ASV, of those represented in the top 15 features. Related to Figure 4.

**Figure S4.**
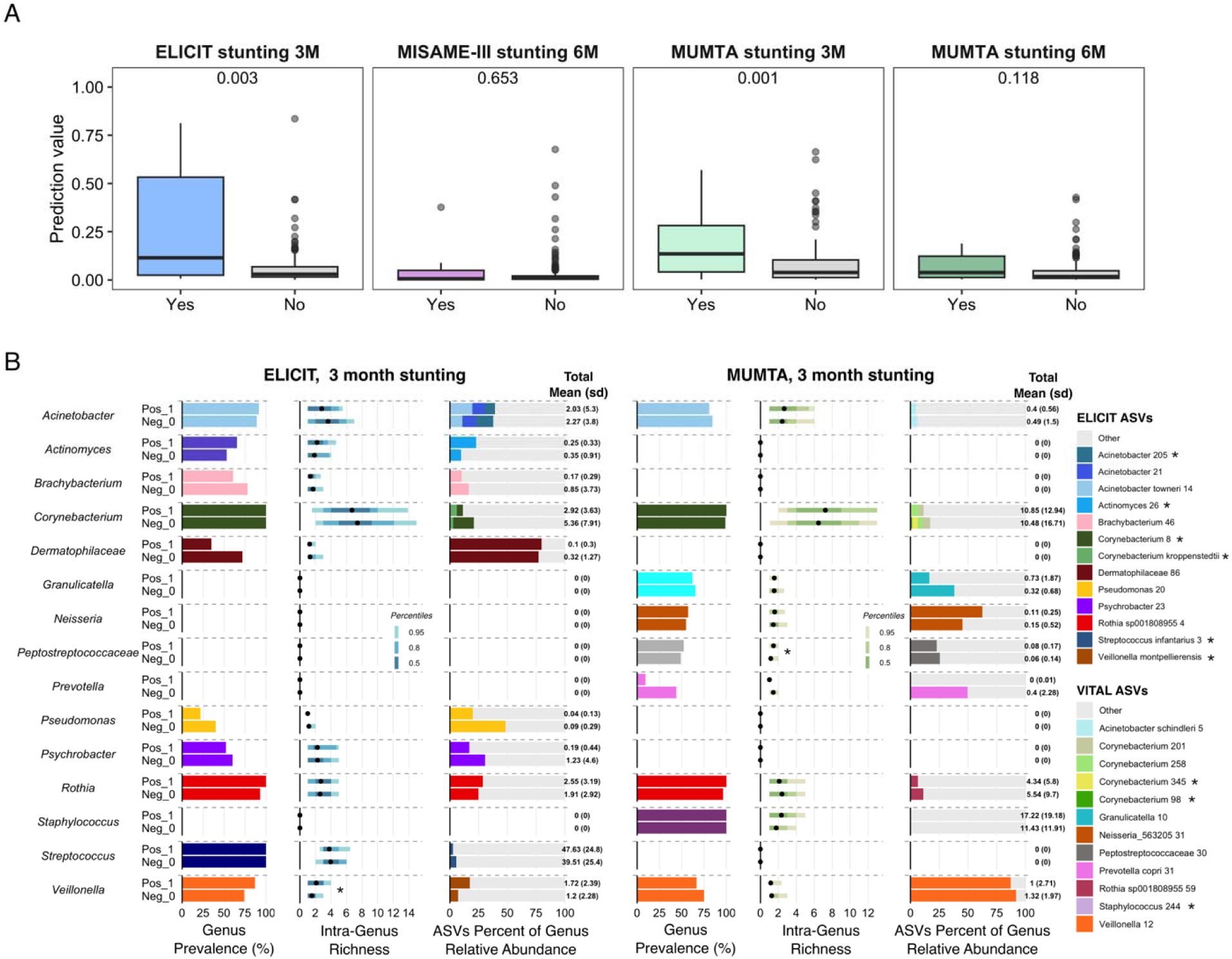
(a) Using xgboost models, human milk microbiota predicted infant stunting at three months with weak-to-moderate accuracy depending on study and growth outcome, based on the following performance metrics: whether the estimated prediction probability was significantly greater in the true case group compared to the control in each study, based on Wilcoxon rank-sum tests, as well as AUROC and AUPRC values at least greater than the prevalence of the rare group (poor growth outcome) in each study (See Figure 5a). (b) Intra-genus richness and composition in the milk of mothers who infants displayed stunted growth at 3 months vs those who did not in ELICIT and MUMTA. Left to right per study: total genus prevalence (left) and within samples that contain the genera, intra-genus richness (the total number of unique ASVs from that genus within a sample) (middle), and the percent of total genus relative abundance contributed by the top 15 ranked ASVs (right). *p<0.05, Wilcoxon rank sum test. Displaying only those genera with >1 unique ASV, of those represented in the top 15 features for a given study (e.g. though *Staphylococcus* is present in ELICIT, it is only among the top 15 most important features for the prediction of stunting in MUMTA, so it is only shown for MUMTA). Related to Figure 5b-c.

**Figure S5.**
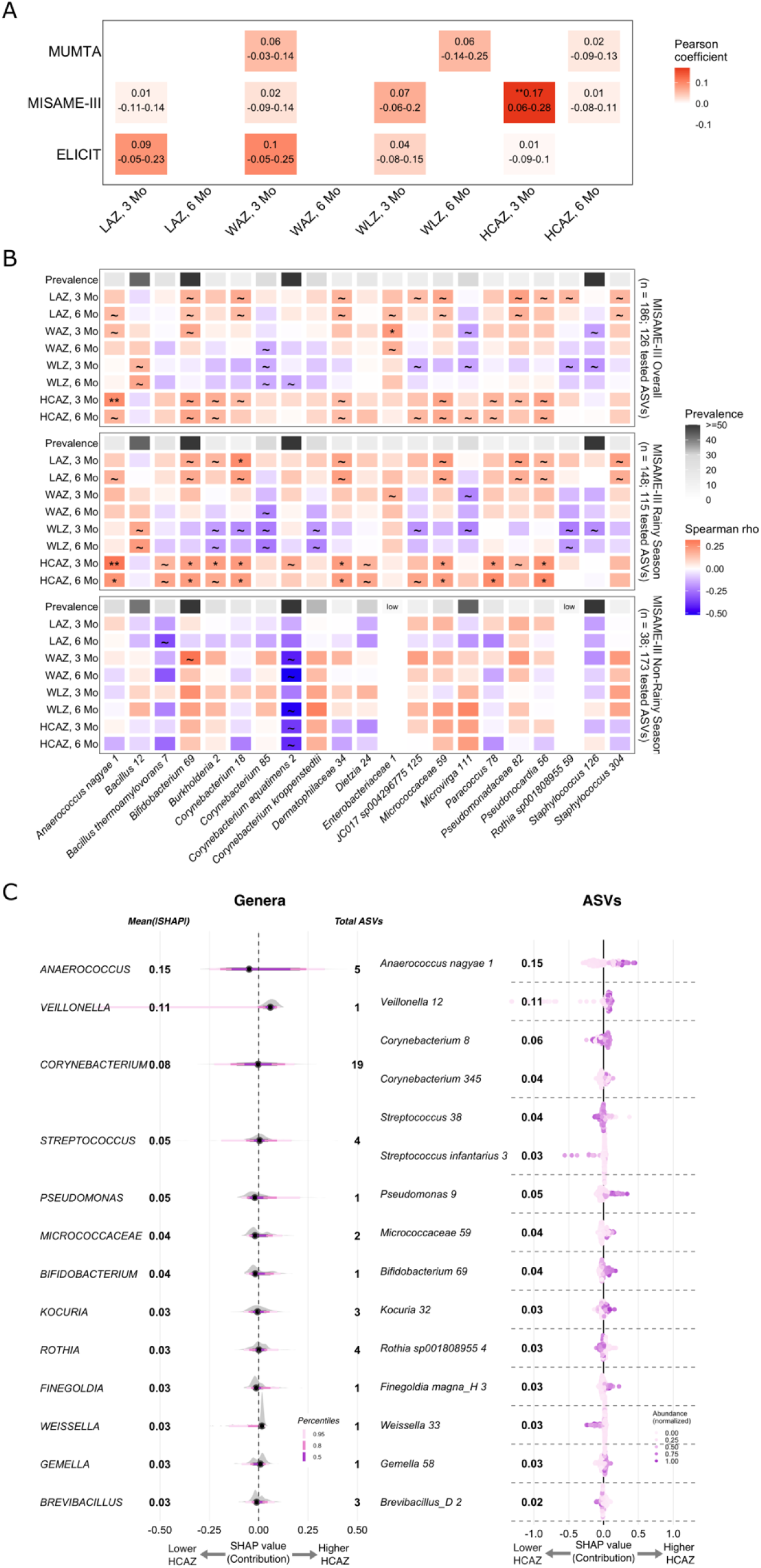
Head circumference z-scores at three months of age displayed a strong association to human milk microbiota in MISAME-III. (a) Using xgboost models, human milk microbiota predicted continuous growth outcomes with low accuracy, with the exception of head circumference z-scores at three months in MISAME-III. Heatmap displays the primary performance metric used based on 10 repeats of a 10-fold cross validated xgboost algorithm: the mean (95% CI) of the Pearson correlation coefficient (*p<0.05, **p<0.001) for the test of whether the outcome value predicted by the xgboost model was positively correlated with the true outcome. White cells indicate mean correlation values ≤0. (b) Spearman correlations of CLR-transformed ASV abundances and infant growth outcomes at three and six months of age in MISAME-III. Heatmap includes ASVs with at least one correlation with a magnitude ≥0.20 and an adjusted p-value <0.3. The top row displays the prevalence of each ASV, with “low” indicating ASVs that were excluded from analysis due to prevalence <0.1. ∼p<0.05; *p(BH)<0.15; **p(BH)<0.05. (c) Feature group (genus) and individual feature (ASV) importance for the top 15 most predictive ASVs (based on the mean of absolute SHAP values) are shown for the model accurately predicting head circumference z-scores at three months in MISAME-III.

**Figure S6.**
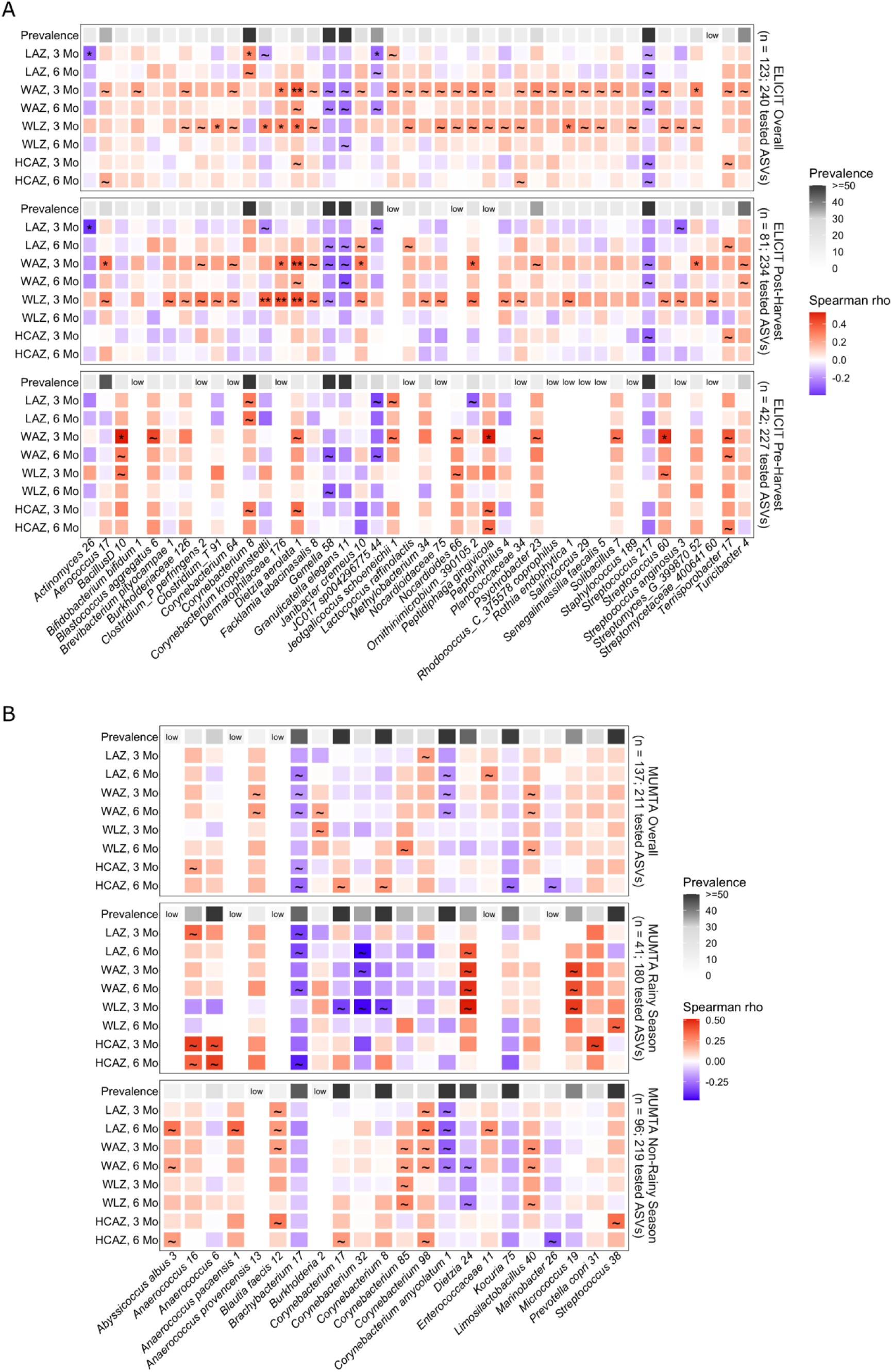
Spearman correlations of CLR-transformed ASV abundances and infant growth outcomes at three and six months of age in (a) ELICIT and (b) MUMTA. The top row displays the prevalence of each ASV, with “low” indicating ASVs that were excluded from analysis due to prevalence <0.1. ∼p<0.05; *p(BH)<0.15; **p(BH)<0.05. (a) The ELICIT heatmap includes ASVs with at least one correlation with a magnitude ≥0.20 and an adjusted p-value <0.3, (b) The MUMTA heatmap includes ASVs with at least one correlation with a magnitude ≥0.20 and an unadjusted p-value <0.01 (only 5 ASVs had an adjusted p-value <0.3).

**Table S1.**
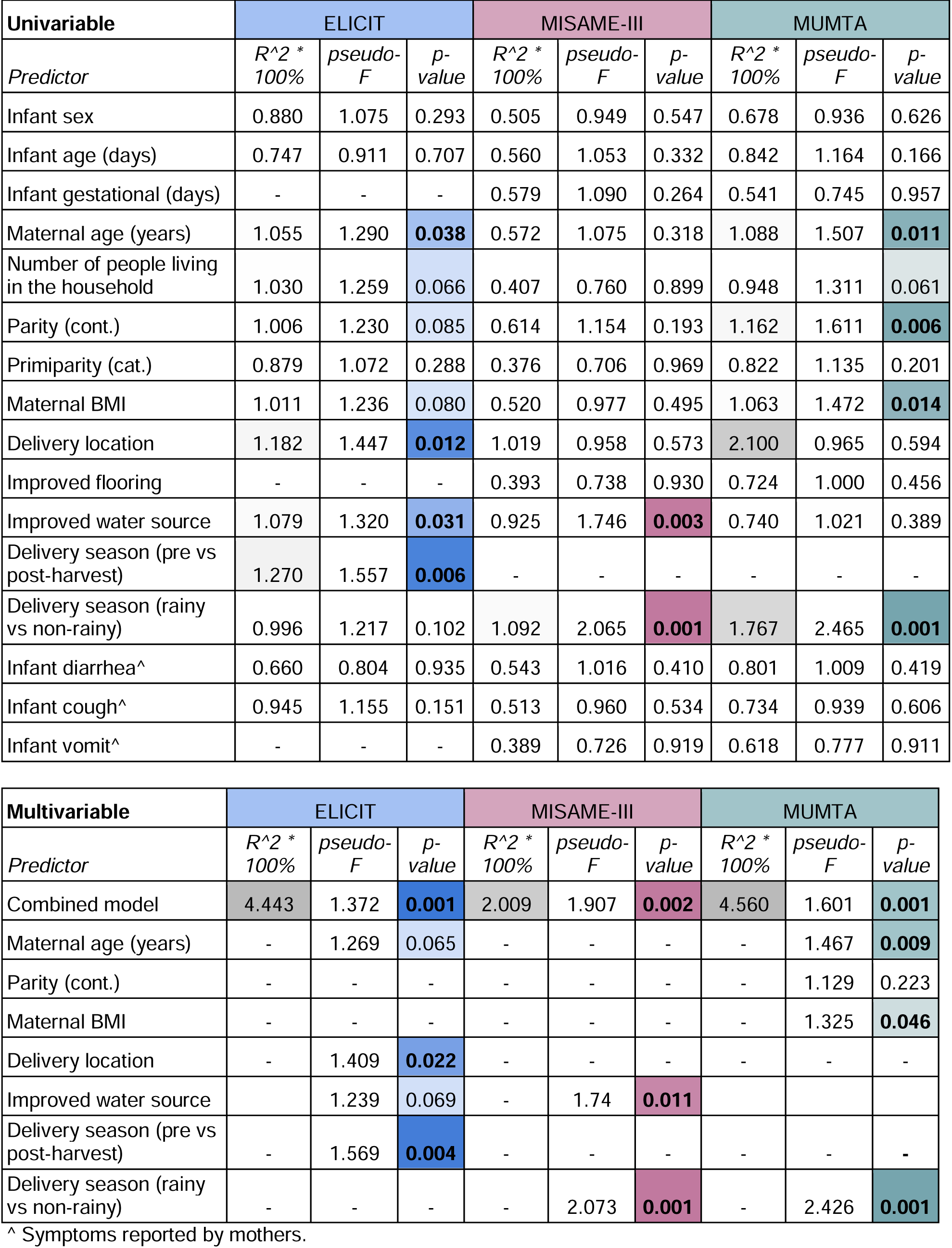
Results of univariable and multivariable redundancy analysis (RDA) of milk bacterial abundance profiles.

**Table S2.** Results of ANCOM-BC2 based on infant delivery season (pre-harvest vs post-harvest) in ELICIT.

| ASV | lfc | se | W | p | q |
| --- | --- | --- | --- | --- | --- |
| *Corynebacterium.314* | 1.307 | 0.175 | 7.487 | 0.000000004 | 0.000000873 |
| *Lautropia mirabilis.1* | 1.481 | 0.178 | 8.326 | 0.000000008 | 0.000000913 |
| *Lactobacillus delbrueckii.5* | 1.283 | 0.196 | 6.555 | 0.000000145 | 0.000010602 |
| *Veillonella_A.12* | -1.540 | 0.279 | -5.517 | 0.000000392 | 0.000017239 |
| *Anaerococcus nagya.13* | 1.513 | 0.228 | 6.639 | 0.000000334 | 0.000017239 |
| *Abiotrophia defectiva.11* | 1.164 | 0.198 | 5.867 | 0.000000492 | 0.000018048 |
| *Lactiplantibacillus.7* | -1.197 | 0.177 | -6.749 | 0.000001123 | 0.000035298 |
| *Moraxella_C_651924.1* | -1.047 | 0.174 | -6.010 | 0.000003963 | 0.000108990 |
| *Unclassified Nocardioideaceae.75* | 1.147 | 0.148 | 7.724 | 0.000009111 | 0.000222720 |
| *Pauljensenia.1* | -1.094 | 0.152 | -7.191 | 0.000010996 | 0.000241910 |
| *Bifidobacterium leopoldii.1* | -0.995 | 0.175 | -5.685 | 0.000014555 | 0.000289930 |
| *Exiguobacterium_A.10* | 0.914 | 0.154 | 5.950 | 0.000015815 | 0.000289930 |
| *Actinomyces.14* | 1.082 | 0.228 | 4.742 | 0.000017386 | 0.000294230 |
| *Corynebacterium propinquum.2* | -0.842 | 0.187 | -4.499 | 0.000034117 | 0.000500380 |
| *Veillonella_A.117* | 0.833 | 0.160 | 5.194 | 0.000032993 | 0.000500380 |
| *Unclassified Dermatophilaceae_390796.176* | 1.062 | 0.168 | 6.318 | 0.000038428 | 0.000528380 |
| *Acetobacter.8* | 1.238 | 0.189 | 6.559 | 0.000040846 | 0.000528590 |
| *Unclassified Moraxellaceae.29* | -0.906 | 0.196 | -4.623 | 0.000059187 | 0.000685320 |
| *Anaerococcus nagya.53* | 1.223 | 0.202 | 6.058 | 0.000056923 | 0.000685320 |
| *Methylobacterium.43* | 0.790 | 0.169 | 4.668 | 0.000063814 | 0.000701950 |
| *Lactococcus_A_346120.2* | 0.978 | 0.216 | 4.538 | 0.000067666 | 0.000705050 |
| *Pseudomonas_A.20* | -0.873 | 0.175 | -4.989 | 0.000070505 | 0.000705050 |
| *Unclassified Weeksellaceae.58* | -0.865 | 0.157 | -5.527 | 0.000074527 | 0.000712860 |
| *Fannyhessea vaginae.1* | -0.854 | 0.163 | -5.247 | 0.000079750 | 0.000731040 |
| *Unclassified Micrococcaceae.33* | 0.914 | 0.185 | 4.940 | 0.000090958 | 0.000800430 |
| *Corynebacterium kroppenstedtii* | 0.899 | 0.204 | 4.416 | 0.000096718 | 0.000804690 |
| *Paracoccus.14* | -0.934 | 0.157 | -5.930 | 0.000098757 | 0.000804690 |
| *Brachybacterium.9* | -0.752 | 0.175 | -4.295 | 0.000144690 | 0.001136820 |
| *Stenotrophomonas_A_615274.14* | 0.792 | 0.152 | 5.196 | 0.000172180 | 0.001262660 |
| *Micrococcus.19* | 0.943 | 0.238 | 3.969 | 0.000170700 | 0.001262660 |
| *Quadrisphaera*.3 | -0.826 | 0.155 | -5.334 | 0.000178310 | 0.001265400 |
| *Staphylococcus*.119 | 0.859 | 0.198 | 4.340 | 0.000192000 | 0.001318960 |
| *Anaerococcus octavius*.19 | 0.766 | 0.172 | 4.456 | 0.000197840 | 0.001318960 |
| *Ligilactobacillus salivarius*.5 | -0.884 | 0.196 | -4.511 | 0.000213070 | 0.001378660 |
| *Brevibacterium pityocampae*.1 | 0.765 | 0.153 | 4.992 | 0.000246620 | 0.001550170 |
| *Terrisporobacter*.17 | 0.664 | 0.157 | 4.241 | 0.000285970 | 0.001747580 |
| *JC017* sp004296775.44 | 0.771 | 0.197 | 3.924 | 0.000309120 | 0.001837990 |
| *Lactobacillus*.31 | -0.838 | 0.185 | -4.542 | 0.000333300 | 0.001929650 |
| *Streptococcus infantarius*.3 | -1.093 | 0.289 | -3.779 | 0.000369160 | 0.002080980 |
| *Chryseobacterium*_796614.47 | 0.815 | 0.167 | 4.880 | 0.000378360 | 0.002080980 |
| *Rothia kristinae* | 0.952 | 0.244 | 3.905 | 0.000425270 | 0.002243650 |
| *Lacticaseibacillus*.4 | 0.844 | 0.184 | 4.580 | 0.000428330 | 0.002243650 |
| *Sphingomonas*_L_486704.93 | 0.705 | 0.152 | 4.631 | 0.000470670 | 0.002408100 |
| *Clostridium*_T.23 | -0.672 | 0.181 | -3.702 | 0.000561340 | 0.002806680 |
| *Streptococcus intermedius*.2 | 0.762 | 0.170 | 4.472 | 0.000629010 | 0.003075170 |
| *Peptidiphaga gingivicola* | 0.798 | 0.172 | 4.631 | 0.000727730 | 0.003480470 |
| *Solibacillus*.7 | 0.612 | 0.151 | 4.046 | 0.000936460 | 0.004383410 |
| Unclassified Pasteurellaceae.1 | -0.761 | 0.208 | -3.660 | 0.000961800 | 0.004408260 |
| *Anaerococcus prevotii*.1 | 0.627 | 0.167 | 3.746 | 0.001056050 | 0.004741460 |
| *Bacillus*_A.12 | 0.792 | 0.223 | 3.556 | 0.001361740 | 0.005874170 |
| *Brachybacterium*.46 | 0.589 | 0.168 | 3.511 | 0.001351510 | 0.005874170 |
| *Corynebacterium*.18 | 0.614 | 0.156 | 3.935 | 0.001496270 | 0.006330360 |
| *Actinomyces massiliensis*.4 | -0.561 | 0.161 | -3.486 | 0.002206070 | 0.009157290 |
| *F0422* sp001553345.42 | 0.867 | 0.272 | 3.190 | 0.002437780 | 0.009931690 |
| *Kocuria*.3 | -0.621 | 0.198 | -3.135 | 0.003054440 | 0.012195190 |
| *Dietzia aerolata*.1 | -0.518 | 0.160 | -3.236 | 0.003104230 | 0.012195190 |
| *Peptoniphilus*_A.10 | 0.633 | 0.200 | 3.170 | 0.003222780 | 0.012438810 |
| *Methylobacterium*.34 | 0.539 | 0.166 | 3.239 | 0.003619860 | 0.013730510 |
| *Peptoniphilus*_A.4 | 0.524 | 0.160 | 3.281 | 0.003739210 | 0.013942820 |
| *Actinomyces graevenitzii*.3 | -0.567 | 0.175 | -3.232 | 0.003994160 | 0.014405180 |
| *Nosocomiicoccus*.3 | -0.547 | 0.156 | -3.497 | 0.003935610 | 0.014405180 |
| Unclassified Enterobacteriaceae_A.34 | 0.759 | 0.251 | 3.022 | 0.004098680 | 0.014543710 |
| *Anaerococcus provencensis*.22 | 0.579 | 0.161 | 3.601 | 0.004165180 | 0.014545080 |
| *Anaerococcus nagyaе.1* | 0.645 | 0.212 | 3.045 | 0.005415090 | 0.018614360 |
| *Jeotgalicoccus_A_310698 schoeneichii.1* | 0.586 | 0.171 | 3.422 | 0.005699510 | 0.019290640 |
| *Dolosigranulum pigrum.19* | -0.601 | 0.213 | -2.826 | 0.006279750 | 0.020836300 |
| *Psychrobacter.38* | 0.546 | 0.172 | 3.170 | 0.006345600 | 0.020836300 |
| *Clostridium_P perfringens.2* | 0.481 | 0.155 | 3.103 | 0.006459630 | 0.020898790 |
| *Corynebacterium glaucum.10* | -0.497 | 0.160 | -3.100 | 0.006873970 | 0.021917000 |
| *Nocardioides_A_392796 glacieisoli* | -0.485 | 0.152 | -3.183 | 0.007204250 | 0.022641930 |
| *Pantoea_A_680069* | 0.600 | 0.207 | 2.899 | 0.008332220 | 0.025818140 |
| *Bifidobacterium bifidum.1* | 0.688 | 0.217 | 3.177 | 0.008814650 | 0.026933660 |
| *Pradoshia* | 0.466 | 0.150 | 3.106 | 0.009990630 | 0.030108760 |
| *Unclassified Planococcaceae.34* | 0.463 | 0.157 | 2.938 | 0.010169310 | 0.030233070 |
| *Unclassified Pseudomonadaceae.81* | 0.504 | 0.171 | 2.941 | 0.011463890 | 0.033627410 |
| *Corynebacterium.278* | 0.452 | 0.151 | 2.990 | 0.012303740 | 0.035278380 |
| *Corynebacterium.345* | 0.459 | 0.166 | 2.764 | 0.012347430 | 0.035278380 |
| *Kocuria.32* | 0.709 | 0.275 | 2.581 | 0.014212300 | 0.040085960 |
| *Unclassified DSM-18226_301387.11* | -0.404 | 0.153 | -2.639 | 0.014979410 | 0.041714810 |
| *Gemella asaccharolytica.1* | 0.414 | 0.150 | 2.754 | 0.015527580 | 0.042700860 |
| *Nocardioides_A_392796.129* | 0.418 | 0.166 | 2.523 | 0.016191760 | 0.043441310 |
| *Rothia aeria.1* | -0.421 | 0.159 | -2.642 | 0.016082940 | 0.043441310 |
| *Anaerococcus nagyaе.48* | 0.475 | 0.176 | 2.698 | 0.016541370 | 0.043844590 |
| *Collinsella.7* | 0.470 | 0.187 | 2.514 | 0.017007870 | 0.044184020 |
| *Neisseria_563205.31* | -0.457 | 0.181 | -2.530 | 0.017075530 | 0.044184020 |
| *Psychrobacter pasteurii.2* | -0.468 | 0.177 | -2.638 | 0.017271940 | 0.044184020 |
| *Granulicatella.10* | 0.482 | 0.201 | 2.396 | 0.019745620 | 0.049931470 |
| *Streptomyces_G_399870.52* | 0.393 | 0.159 | 2.475 | 0.020460420 | 0.050625090 |
| *Weissella_A_338544.33* | 0.467 | 0.192 | 2.431 | 0.020480150 | 0.050625090 |
| *Anaerococcus.84* | -0.425 | 0.175 | -2.428 | 0.021635680 | 0.052887220 |
| *Bradyrhizobium* | 0.397 | 0.151 | 2.639 | 0.023045030 | 0.055107690 |
| *Blastococcus.2* | -0.428 | 0.169 | -2.536 | 0.022829590 | 0.055107690 |
| *Enteractinococcus fodinae.2* | 0.404 | 0.160 | 2.521 | 0.024441730 | 0.057819150 |
| *Corynebacterium.64* | 0.364 | 0.150 | 2.424 | 0.027570580 | 0.063351240 |
| *Paracoccus.78* | 0.448 | 0.191 | 2.347 | 0.027884120 | 0.063351240 |
| *Leuconostoc_B falkenbergense.3* | 0.462 | 0.198 | 2.334 | 0.027932140 | 0.063351240 |
| *Escherichia_710834.34* | -0.607 | 0.269 | -2.260 | 0.027652580 | 0.063351240 |
| *Lentilactobacillus.8* | 0.403 | 0.166 | 2.425 | 0.029433490 | 0.066075170 |
| *Corynebacterium.304* | -0.361 | 0.152 | -2.368 | 0.034067740 | 0.075076530 |
| *Acinetobacter.205* | 0.511 | 0.236 | 2.167 | 0.034125700 | 0.075076530 |
| *Unclassified Dermatophilaceae_390796.34* | 0.405 | 0.177 | 2.284 | 0.034707310 | 0.075600080 |
| *Corynebacterium.8* | 0.635 | 0.298 | 2.132 | 0.036048640 | 0.077475160 |
| *Rothia sp001808955.33* | -0.450 | 0.208 | -2.163 | 0.036272460 | 0.077475160 |
| *Unclassified Enterococcaceae.11* | 0.474 | 0.213 | 2.228 | 0.037517480 | 0.079329470 |
| *JC017 sp004296775.125* | 0.452 | 0.214 | 2.118 | 0.037861790 | 0.079329470 |
| *Ochrobactrum_A_499024 pseudogrignonensis.1* | -0.351 | 0.157 | -2.235 | 0.043605690 | 0.090502380 |
| *Rhodococcus_C_375578 coprophilus* | 0.346 | 0.156 | 2.223 | 0.044556100 | 0.091610680 |
| *Corynebacterium.201* | 0.631 | 0.314 | 2.008 | 0.047227860 | 0.096204910 |
| *Rothia sp001808955.4* | 0.443 | 0.217 | 2.045 | 0.047685210 | 0.096245390 |
| *Psychrobacter.23* | -0.337 | 0.165 | -2.037 | 0.049760070 | 0.099520130 |
| *Paracoccus acridae.2* | 0.342 | 0.171 | 1.996 | 0.056077750 | 0.111145090 |
| *Kytococcus sedentarius.7* | -0.312 | 0.156 | -1.998 | 0.057696690 | 0.113332790 |
| *Psychrobacter sanguinis_A* | 0.387 | 0.198 | 1.952 | 0.060695640 | 0.118168500 |
| *Corynebacterium aurimucosum.7* | 0.359 | 0.182 | 1.967 | 0.062483210 | 0.120581640 |
| *Actinomyces timonensis.1* | -0.338 | 0.163 | -2.067 | 0.063135860 | 0.120781650 |
| *Dietzia cinnamomea.11* | 0.365 | 0.194 | 1.881 | 0.066330570 | 0.123667170 |
| *Corynebacterium sp001767255.1* | 0.311 | 0.152 | 2.039 | 0.066261930 | 0.123667170 |
| *Neobacillus.24* | -0.317 | 0.168 | -1.891 | 0.065742330 | 0.123667170 |
| *Streptococcus.60* | -0.363 | 0.184 | -1.980 | 0.067731240 | 0.125217410 |
| *Streptococcus sanguinis_H.4* | 0.406 | 0.224 | 1.811 | 0.079274060 | 0.145335780 |
| *Streptococcus.217* | -0.597 | 0.337 | -1.768 | 0.080021730 | 0.145494060 |
| *Unclassified Burkholderiaceae_A_592522.126* | 0.285 | 0.152 | 1.872 | 0.080882640 | 0.145853930 |
| *Georgenia wutianyii* | -0.320 | 0.177 | -1.809 | 0.085526970 | 0.152571910 |
| *Bifidobacterium_388775.69* | -0.607 | 0.350 | -1.736 | 0.085995080 | 0.152571910 |
| *Anaerococcus vaginimassiliensis.5* | 0.341 | 0.195 | 1.751 | 0.088181260 | 0.153967270 |
| *Peptostreptococcus stomatis.2* | 0.308 | 0.168 | 1.836 | 0.087683140 | 0.153967270 |
| *Kocuria.75* | -0.413 | 0.240 | -1.717 | 0.090469380 | 0.156718620 |
| *Aeromonas.1* | -0.356 | 0.207 | -1.718 | 0.093605610 | 0.160884650 |
| *Pseudomonas_A.9* | 0.292 | 0.169 | 1.729 | 0.095308540 | 0.162541700 |
| *Clostridium_T.91* | 0.274 | 0.159 | 1.718 | 0.099832910 | 0.168948010 |
| *Facklamia_A_322620 tabacinasalis.8* | -0.265 | 0.157 | -1.683 | 0.105817120 | 0.177708140 |
| *Dietzia.24* | -0.330 | 0.214 | -1.541 | 0.127367280 | 0.212278800 |
| *Acinetobacter.115* | -0.355 | 0.235 | -1.513 | 0.137882300 | 0.228075980 |
| *Massilia.32* | 0.251 | 0.161 | 1.558 | 0.140174850 | 0.230137810 |
| *Kocuria.29* | -0.386 | 0.263 | -1.465 | 0.146993820 | 0.239545490 |
| *Staphylococcus.304* | 0.274 | 0.183 | 1.495 | 0.149027010 | 0.241073110 |
| *Blastococcus aggregatus.6* | -0.259 | 0.176 | -1.475 | 0.153147290 | 0.245929950 |
| *Dietzia lutea* | -0.262 | 0.183 | -1.431 | 0.157575190 | 0.251206830 |
| *Acinetobacter schindleri.5* | 0.306 | 0.217 | 1.414 | 0.164515390 | 0.260384070 |
| *Streptococcus.38* | -0.499 | 0.372 | -1.344 | 0.181467450 | 0.281378600 |
| *Corynebacterium.118* | -0.218 | 0.154 | -1.411 | 0.181617100 | 0.281378600 |
| *Unclassified Peptostreptococcaceae_256921.30* | -0.243 | 0.177 | -1.370 | 0.179953050 | 0.281378600 |
| *Corynebacterium.17* | 0.246 | 0.182 | 1.350 | 0.190859050 | 0.293629300 |
| *Pauljensenia sp000308055.3* | -0.246 | 0.182 | -1.350 | 0.193876880 | 0.296200780 |
| *Brevibacterium.24* | 0.208 | 0.160 | 1.301 | 0.211741590 | 0.321263100 |
| *Unclassified Enterobacteriaceae_A.67* | 0.278 | 0.221 | 1.254 | 0.218231630 | 0.328842180 |
| *Corynebacterium.71* | -0.190 | 0.163 | -1.170 | 0.257261030 | 0.374817390 |
| *Bifidobacterium vaginale* | -0.238 | 0.205 | -1.163 | 0.254471890 | 0.374817390 |
| *Nocardioides_A_392796.221* | 0.194 | 0.162 | 1.198 | 0.256244750 | 0.374817390 |
| *Bacillus_BD.10* | -0.181 | 0.154 | -1.172 | 0.254335520 | 0.374817390 |
| *Peptostreptococcus anaerobius.3* | 0.199 | 0.167 | 1.192 | 0.254445900 | 0.374817390 |
| *Paracoccus carotinifaciens.12* | 0.294 | 0.259 | 1.133 | 0.262247480 | 0.379568720 |
| *Niallia oryzisoli.17* | -0.208 | 0.189 | -1.104 | 0.273293830 | 0.392971510 |
| *Staphylococcus.126* | 0.274 | 0.253 | 1.082 | 0.281268620 | 0.399219980 |
| *Actinomyces.26* | -0.190 | 0.171 | -1.112 | 0.280947320 | 0.399219980 |
| *Citricoccus sp000224415.5* | 0.162 | 0.152 | 1.066 | 0.294874420 | 0.415848530 |
| *Finegoldia magna_H.3* | 0.198 | 0.188 | 1.056 | 0.296953580 | 0.416113290 |
| *Janibacter_A_390549 cremeus.10* | -0.184 | 0.174 | -1.055 | 0.300322720 | 0.418170880 |
| *Glutamicibacter.5* | -0.173 | 0.167 | -1.034 | 0.313046070 | 0.433145510 |
| *Corynebacterium amycolatum_A.1* | 0.293 | 0.293 | 0.999 | 0.320079150 | 0.440108830 |
| *Rothia sp001808955.59* | -0.172 | 0.180 | -0.955 | 0.347269590 | 0.474529880 |
| *Nocardioides_A_392796.66* | -0.143 | 0.154 | -0.928 | 0.373237470 | 0.506865700 |
| *Corynebacterium stationis*.2 | -0.139 | 0.153 | -0.908 | 0.376063570 | 0.507570460 |
| *Neisseria*_563205.32 | 0.187 | 0.216 | 0.866 | 0.391786850 | 0.525567720 |
| *Acinetobacter*.21 | -0.182 | 0.216 | -0.844 | 0.403045890 | 0.536878950 |
| *Staphylococcus*.52 | 0.144 | 0.169 | 0.853 | 0.405099570 | 0.536878950 |
| *Arthrobacter*_D.1 | 0.126 | 0.156 | 0.813 | 0.424984580 | 0.559859920 |
| *Ornithinimicrobium*_390105.2 | 0.131 | 0.170 | 0.769 | 0.449214440 | 0.586927320 |
| *Aerococcus*.17 | -0.159 | 0.209 | -0.761 | 0.450866900 | 0.586927320 |
| *Haemophilus*_D_735815.20 | -0.141 | 0.188 | -0.748 | 0.458459410 | 0.593300410 |
| Unclassified *Enterobacteriaceae*_A.15 | 0.143 | 0.190 | 0.756 | 0.465306350 | 0.598639740 |
| *Corynebacterium*.258 | 0.124 | 0.173 | 0.720 | 0.476603700 | 0.609609390 |
| *Streptococcus*.341 | 0.224 | 0.332 | 0.674 | 0.501845250 | 0.638184710 |
| Unclassified *Peptostreptococcaceae*_256921.26 | 0.120 | 0.180 | 0.667 | 0.509012250 | 0.643578700 |
| *Corynebacterium maris*.5 | -0.118 | 0.193 | -0.612 | 0.544775530 | 0.680969410 |
| *Acinetobacter*.124 | 0.117 | 0.191 | 0.615 | 0.542705560 | 0.680969410 |
| *Peptococcus_niger* | 0.098 | 0.165 | 0.596 | 0.560375810 | 0.688267410 |
| *Clostridioides*_A_difficile.5 | 0.105 | 0.184 | 0.569 | 0.572513340 | 0.688267410 |
| *Staphylococcus*.189 | -0.100 | 0.170 | -0.589 | 0.566979970 | 0.688267410 |
| *Prevotella copri*.31 | 0.089 | 0.151 | 0.584 | 0.566247970 | 0.688267410 |
| *Limosilactobacillus*.34 | -0.107 | 0.184 | -0.581 | 0.568525130 | 0.688267410 |
| *Microvirga*.111 | 0.091 | 0.156 | 0.580 | 0.568939410 | 0.688267410 |
| *Salinicoccus*.28 | -0.101 | 0.177 | -0.572 | 0.570488750 | 0.688267410 |
| *Corynebacterium durum*.2 | -0.091 | 0.164 | -0.556 | 0.585345560 | 0.699869690 |
| *Veillonella*_A.91 | 0.086 | 0.163 | 0.525 | 0.605458130 | 0.720004270 |
| Unclassified *Enterobacteriaceae*_A.1 | -0.085 | 0.163 | -0.519 | 0.610955970 | 0.722636100 |
| *Corynebacterium*.85 | 0.103 | 0.212 | 0.488 | 0.627586020 | 0.726678550 |
| *Corynebacterium*.234 | -0.085 | 0.174 | -0.492 | 0.626331180 | 0.726678550 |
| *Pseudonocardia*.56 | -0.076 | 0.152 | -0.499 | 0.623537820 | 0.726678550 |
| *Rothia dentocariosa*.4 | -0.080 | 0.162 | -0.492 | 0.626640920 | 0.726678550 |
| *Alloprevotella*_sp900095835.14 | -0.093 | 0.200 | -0.467 | 0.646634050 | 0.744814090 |
| *Pauljensenia*.35 | 0.123 | 0.271 | 0.454 | 0.651749860 | 0.746796710 |
| *Rothia terrae*.8 | 0.098 | 0.222 | 0.442 | 0.661383880 | 0.753909080 |
| *Piscicoccus intestinalis*.4 | 0.074 | 0.178 | 0.414 | 0.682349920 | 0.762015140 |
| *Anaerococcus vaginalis*.2 | -0.066 | 0.155 | -0.423 | 0.680433730 | 0.762015140 |
| *Psychrobacter sanguinis*_A.2 | -0.092 | 0.219 | -0.420 | 0.677807650 | 0.762015140 |
| *Priestia*.20 | -0.067 | 0.159 | -0.419 | 0.681036050 | 0.762015140 |
| *Acinetobacter*.16 | 0.115 | 0.296 | 0.388 | 0.699313000 | 0.769244300 |
| *Unclassified Micrococcaceae*.59 | 0.073 | 0.187 | 0.393 | 0.695556310 | 0.769244300 |
| *Rothia* sp001808955.60 | 0.082 | 0.208 | 0.392 | 0.696582960 | 0.769244300 |
| *Peptoniphilus*_C.1 | 0.063 | 0.166 | 0.378 | 0.708367640 | 0.772391480 |
| *Salinicoccus*.7 | 0.062 | 0.165 | 0.379 | 0.709195810 | 0.772391480 |
| *Anaerococcus prevotii*.8 | 0.064 | 0.176 | 0.366 | 0.716809580 | 0.776837970 |
| *Turicibacter*.4 | 0.060 | 0.178 | 0.336 | 0.738297510 | 0.793166670 |
| *Unclassified Dermatophilaceae*_390796.86 | -0.072 | 0.217 | -0.334 | 0.739087120 | 0.793166670 |
| *Psychrobacter*.8 | -0.055 | 0.179 | -0.309 | 0.760013430 | 0.811664830 |
| *Kocuria palustris*.9 | -0.099 | 0.328 | -0.301 | 0.763780290 | 0.811747170 |
| *Granulicatella elegans*.11 | 0.079 | 0.288 | 0.275 | 0.784319730 | 0.829568940 |
| *Romboutsia*_B ilealis.1 | -0.051 | 0.209 | -0.243 | 0.808767410 | 0.851334110 |
| *Microvirga*.131 | -0.035 | 0.150 | -0.235 | 0.818683600 | 0.857668530 |
| *Leuconostoc*_B.8 | 0.045 | 0.216 | 0.209 | 0.835298970 | 0.870927840 |
| *Jeotgalicoccus*_A_310962.2 | -0.032 | 0.168 | -0.189 | 0.852186200 | 0.884344170 |
| *Acinetobacter*.203 | 0.027 | 0.160 | 0.167 | 0.869146380 | 0.897709880 |
| *Chryseomicrobium excrementi*.2 | -0.024 | 0.164 | -0.146 | 0.886392530 | 0.907006310 |
| *Acinetobacter townneri*_A.14 | 0.028 | 0.197 | 0.144 | 0.885849720 | 0.907006310 |
| *Sarcina ventriculi*.12 | 0.020 | 0.159 | 0.128 | 0.899698200 | 0.914122870 |
| *Corynebacterium*.429 | 0.021 | 0.167 | 0.124 | 0.901657560 | 0.914122870 |
| *Brachybacterium*.17 | 0.025 | 0.245 | 0.103 | 0.918525210 | 0.926952050 |
| *Gemella*.58 | 0.015 | 0.339 | 0.045 | 0.963823690 | 0.968224710 |
| *Limosilactobacillus fermentum*.5 | 0.008 | 0.211 | 0.038 | 0.970119090 | 0.970119090 |

**Table S3.** Results of ANCOM-BC2 based on delivery season (rainy season vs summer season) in MISAME-III.

| ASV | lfc | se | W | p | q |
| --- | --- | --- | --- | --- | --- |
| *Rothia* sp001808955.33 | -2.117 | 0.138 | -15.305 | 0.00000000 | 0.00000000 |
| *Bosea*.27 | 1.464 | 0.102 | 14.389 | 0.00000000 | 0.00000000 |
| *Pseudomonas*_E_648040 *alcaligenes* | 1.568 | 0.105 | 14.963 | 0.00000000 | 0.00000000 |
| *Brevundimonas*.18 | 1.114 | 0.124 | 9.015 | 0.00000000 | 0.00000000 |
| *Aerococcus*.17 | 1.126 | 0.090 | 12.520 | 0.00000000 | 0.00000000 |
| Unclassified *Nocardoidaceae*.82 | 1.237 | 0.091 | 13.517 | 0.00000000 | 0.00000000 |
| Unclassified *Pseudomonadaceae*.82 | 1.476 | 0.135 | 10.962 | 0.00000000 | 0.00000001 |
| *Pseudomonas*_E_650326 *otitidis* | 1.022 | 0.118 | 8.677 | 0.00000001 | 0.00000014 |
| *Corynebacterium*.85 | 0.755 | 0.113 | 6.662 | 0.00000002 | 0.00000027 |
| *Staphylococcus*.304 | -1.240 | 0.155 | -8.002 | 0.00000002 | 0.00000027 |
| *Piscicoccus intestinalis*.4 | 0.979 | 0.147 | 6.667 | 0.00000007 | 0.00000070 |
| *Acinetobacter*.43 | -1.306 | 0.198 | -6.602 | 0.00000008 | 0.00000070 |
| Unclassified *Mycobacteriaceae*.15 | -0.855 | 0.104 | -8.251 | 0.00000011 | 0.00000090 |
| *Granulicatella*.10 | 0.966 | 0.132 | 7.311 | 0.00000019 | 0.00000154 |
| *Anaerococcus nagyae*.13 | -0.870 | 0.120 | -7.267 | 0.00000021 | 0.00000158 |
| *Mycobacterium*.68 | 0.792 | 0.099 | 8.042 | 0.00000023 | 0.00000158 |
| Unclassified *Dermatophilaceae*_390796.34 | 0.647 | 0.093 | 6.982 | 0.00000041 | 0.00000266 |
| *Stenotrophomonas*_A_615274.14 | 0.851 | 0.141 | 6.020 | 0.00000044 | 0.00000273 |
| *Anaerococcus provencensis*.13 | 0.900 | 0.119 | 7.589 | 0.00000051 | 0.00000301 |
| *Acinetobacter*.124 | 0.968 | 0.176 | 5.488 | 0.00000059 | 0.00000324 |
| *Aquabacterium*_B_592457 *olei*.2 | 0.793 | 0.118 | 6.709 | 0.00000061 | 0.00000324 |
| *Rhodococcus*_C_375578.9 | 0.731 | 0.111 | 6.593 | 0.00000158 | 0.00000796 |
| *Blastococcus aggregatus*.6 | 0.646 | 0.112 | 5.778 | 0.00000260 | 0.00001254 |
| Unclassified *Burkholderiaceae*_A_592522.27 | -0.580 | 0.095 | -6.075 | 0.00000339 | 0.00001568 |
| *Bacillus*_A.12 | 1.301 | 0.264 | 4.926 | 0.00000452 | 0.00002006 |
| *Paracoccus acridae*.2 | 0.586 | 0.095 | 6.141 | 0.00000666 | 0.00002845 |
| *Clostridium*_T.23 | 0.563 | 0.094 | 5.966 | 0.00000780 | 0.00003207 |
| *Staphylococcus*.189 | -1.013 | 0.184 | -5.495 | 0.00001194 | 0.00004732 |
| *Paracoccus*.78 | 0.579 | 0.102 | 5.650 | 0.00001315 | 0.00005032 |
| *Niallia oryzisoli*.17 | -0.483 | 0.090 | -5.366 | 0.00001650 | 0.00006104 |
| *Corynebacterium aurimucosum*.7 | -0.699 | 0.155 | -4.506 | 0.00003778 | 0.00013526 |
| *Bradyrhizobium* | -0.476 | 0.097 | -4.892 | 0.00004472 | 0.00015512 |
| *Finnegoldia magna*_H.3 | -0.610 | 0.135 | -4.525 | 0.00007031 | 0.00023649 |
| *Acinetobacter baumannii* | 1.038 | 0.255 | 4.075 | 0.00010987 | 0.00035871 |
| *Romboutsia_B ilealis*.1 | 0.545 | 0.132 | 4.145 | 0.00012578 | 0.00039890 |
| *Limosilactobacillus fermentum*.5 | -0.578 | 0.146 | -3.964 | 0.00026108 | 0.00080499 |
| JC017 sp004296775.125 | 0.400 | 0.101 | 3.961 | 0.00061897 | 0.00185690 |
| *Escherichia_710834*.34 | -0.707 | 0.184 | -3.833 | 0.00068662 | 0.00200566 |
| *Bacillus_P_294101*.28 | 0.489 | 0.136 | 3.598 | 0.00076938 | 0.00218978 |
| *Nocardioides_A_392796*.129 | 0.395 | 0.102 | 3.883 | 0.00100055 | 0.00277653 |
| *Moraxella_A_651148 atlantae*.20 | -0.429 | 0.118 | -3.640 | 0.00105321 | 0.00282085 |
| Unclassified Moraxellaceae.29 | 0.461 | 0.131 | 3.509 | 0.00106735 | 0.00282085 |
| Unclassified Enterobacteriaceae_A.34 | 0.867 | 0.257 | 3.374 | 0.00110289 | 0.00284699 |
| *Stenotrophomonas_A_615274*.5 | 0.471 | 0.129 | 3.656 | 0.00119070 | 0.00300382 |
| *Agrobacterium*.1 | 0.427 | 0.127 | 3.368 | 0.00203707 | 0.00502478 |
| *Kocuria*.29 | -0.418 | 0.118 | -3.539 | 0.00251911 | 0.00607872 |
| *Streptococcus infantarius*.3 | 0.559 | 0.176 | 3.180 | 0.00307906 | 0.00727182 |
| *Rothia terrae*.8 | 0.396 | 0.138 | 2.880 | 0.00704327 | 0.01628756 |
| *Dietzia*.24 | 0.312 | 0.115 | 2.717 | 0.01054001 | 0.02387636 |
| *Massilia*.19 | -0.360 | 0.131 | -2.745 | 0.01126646 | 0.02501154 |
| *Turicibacter*.4 | 0.306 | 0.114 | 2.681 | 0.01259007 | 0.02687497 |
| Unclassified Planococcaceae.34 | 0.289 | 0.105 | 2.746 | 0.01244334 | 0.02687497 |
| *Aeromonas*.1 | -0.328 | 0.125 | -2.634 | 0.01426519 | 0.02962205 |
| *Weissella_A_338544*.33 | 0.357 | 0.139 | 2.571 | 0.01441073 | 0.02962205 |
| *Bacillus_J thermoamylovorans*.7 | 0.252 | 0.099 | 2.535 | 0.01547291 | 0.03122714 |
| *Corynebacterium*.107 | 0.331 | 0.131 | 2.521 | 0.01985876 | 0.03936290 |
| *Corynebacterium*.18 | 0.242 | 0.098 | 2.479 | 0.02175820 | 0.04237124 |
| *Corynebacterium kroppenstedtii* | 0.299 | 0.129 | 2.324 | 0.02420472 | 0.04632282 |
| Unclassified Micrococcaceae.33 | 0.311 | 0.131 | 2.370 | 0.02855609 | 0.05372418 |
| *Dietzia cinnamnea*.11 | 0.356 | 0.164 | 2.178 | 0.03255118 | 0.06021969 |
| *Pseudomonas_A*.9 | 0.223 | 0.104 | 2.134 | 0.04375068 | 0.07961190 |
| *Veillonella_A*.12 | 0.518 | 0.257 | 2.016 | 0.04618915 | 0.08269349 |
| Unclassified Pseudomonadaceae.70 | -0.276 | 0.137 | -2.020 | 0.05241812 | 0.09235573 |
| *Burkholderia*.2 | -0.200 | 0.100 | -1.993 | 0.05610176 | 0.09730148 |
| *Corynebacterium*.172 | 0.225 | 0.112 | 2.016 | 0.05748180 | 0.09816122 |
| *Anaerococcus*.6 | -0.231 | 0.120 | -1.924 | 0.06634027 | 0.10996809 |
| *Acinetobacter*.16 | -0.276 | 0.145 | -1.910 | 0.06637713 | 0.10996809 |
| *Corynebacterium*.8 | -0.511 | 0.289 | -1.767 | 0.07926398 | 0.12928254 |
| *Streptococcus*.217 | 0.481 | 0.272 | 1.768 | 0.08036482 | 0.12928254 |
| *Methylobacterium*.34 | -0.294 | 0.164 | -1.797 | 0.08273808 | 0.13119895 |
| *Corynebacterium*.32 | -0.193 | 0.108 | -1.786 | 0.09201889 | 0.14386052 |
| *Corynebacterium aquatimens*.2 | -0.360 | 0.226 | -1.595 | 0.11352584 | 0.17501901 |
| *Unclassified Burkholderiaceae_A_592522*.133 | 0.172 | 0.104 | 1.652 | 0.11581705 | 0.17610538 |
| *Cytobacillus_298193*.4 | -0.212 | 0.134 | -1.579 | 0.12452625 | 0.18678938 |
| *Micrococcus*.19 | -0.264 | 0.173 | -1.520 | 0.13184547 | 0.19513130 |
| *Corynebacterium riegelii*.3 | 0.168 | 0.111 | 1.516 | 0.13620188 | 0.19892643 |
| *Moraxella_A_651148 atlantae*.1 | -0.214 | 0.142 | -1.506 | 0.13846884 | 0.19961092 |
| *Exiguobacterium_A*.10 | -0.220 | 0.154 | -1.429 | 0.16580331 | 0.23595086 |
| *Anaerococcus nagyae*.1 | -0.152 | 0.122 | -1.246 | 0.22187264 | 0.31174511 |
| *Corynebacterium*.17 | -0.302 | 0.271 | -1.113 | 0.26783140 | 0.36704030 |
| *Brevibacillus_D*.2 | 0.155 | 0.138 | 1.128 | 0.26804910 | 0.36704030 |
| *Priestia*.20 | 0.261 | 0.235 | 1.110 | 0.27114689 | 0.36704030 |
| *Staphylococcus*.126 | -0.268 | 0.269 | -0.997 | 0.32020718 | 0.42313092 |
| *Gemella*.58 | -0.252 | 0.252 | -1.003 | 0.31815206 | 0.42313092 |
| *Rothia sp001808955*.4 | 0.211 | 0.217 | 0.972 | 0.33366191 | 0.43572320 |
| *Brevibacillus_D*.34 | 0.086 | 0.090 | 0.954 | 0.35219345 | 0.45457526 |
| *Corynebacterium glaucum*.10 | -0.089 | 0.110 | -0.804 | 0.42640502 | 0.54403399 |
| *Neobacillus*.24 | 0.110 | 0.139 | 0.789 | 0.43323048 | 0.54646117 |
| *Kocuria*.32 | 0.194 | 0.292 | 0.665 | 0.50745031 | 0.63288747 |
| *Corynebacterium*.258 | 0.109 | 0.184 | 0.594 | 0.55353855 | 0.67519538 |
| *Kocuria*.75 | 0.091 | 0.153 | 0.596 | 0.55277571 | 0.67519538 |
| *Streptococcus*.341 | -0.162 | 0.346 | -0.467 | 0.64109380 | 0.77349360 |
| *Moraxella_A_651148 atlantae*.5 | -0.078 | 0.176 | -0.441 | 0.66216740 | 0.79032884 |
| *Pseudonocardia*.56 | -0.038 | 0.090 | -0.418 | 0.67923439 | 0.80207466 |
| *Streptococcus*.38 | 0.116 | 0.349 | 0.331 | 0.74070902 | 0.83327279 |
| *JC017 sp004296775*.44 | 0.051 | 0.166 | 0.304 | 0.76259810 | 0.83327279 |
| *Corynebacterium.110* | -0.030 | 0.092 | -0.330 | 0.74543152 | 0.83327279 |
| *Corynebacterium.201* | -0.068 | 0.213 | -0.318 | 0.75157434 | 0.83327279 |
| *Methylobacterium.43* | 0.036 | 0.111 | 0.328 | 0.74644459 | 0.83327279 |
| *Unclassified Micrococcaceae.59* | 0.038 | 0.125 | 0.303 | 0.76403581 | 0.83327279 |
| *Comamonas_F_589250.14* | 0.044 | 0.135 | 0.330 | 0.74309398 | 0.83327279 |
| *Paracoccus.126* | 0.028 | 0.093 | 0.302 | 0.76571013 | 0.83327279 |
| *Corynebacterium.345* | -0.036 | 0.136 | -0.265 | 0.79257513 | 0.85413437 |
| *Bacillus_P_294101.36* | 0.044 | 0.187 | 0.238 | 0.81259498 | 0.86728887 |
| *Brevibacillus_D.24* | -0.029 | 0.202 | -0.143 | 0.88723736 | 0.92955896 |
| *Microvirga.111* | 0.021 | 0.147 | 0.142 | 0.88768693 | 0.92955896 |
| *Anaerococcus.16* | -0.015 | 0.175 | -0.088 | 0.93010756 | 0.96487794 |
| *Collinsella.7* | -0.007 | 0.116 | -0.060 | 0.95233254 | 0.97878622 |
| *Corynebacterium mycetoides.2* | -0.001 | 0.102 | -0.008 | 0.99351876 | 0.99351876 |
| *Bifidobacterium_388775.69* | -0.002 | 0.257 | -0.009 | 0.99296824 | 0.99351876 |
| *Corynebacterium amycolatum_A.1* | 0.005 | 0.233 | 0.020 | 0.98411761 | 0.99351876 |

**Table S4.** Results of ANCOM-BC2 based on delivery season (rainy season vs non-rainy season) in MUMTA.

| ASV | lfc | se | W | p | q |
| --- | --- | --- | --- | --- | --- |
| *Corynebacterium.8* | 2.310 | 0.344 | 6.706 | 0.00000000 | 0.00000012 |
| *Rothia sp001808955.59* | -1.478 | 0.201 | -7.359 | 0.00000001 | 0.00000046 |
| *Alkalibacterium.1* | -1.364 | 0.176 | -7.751 | 0.00000001 | 0.00000046 |
| *Corynebacterium.17* | 1.796 | 0.314 | 5.723 | 0.00000015 | 0.00000656 |
| *Anaerococcus.16* | 1.578 | 0.220 | 7.166 | 0.00000021 | 0.00000754 |
| *Anaerococcus.6* | 1.447 | 0.246 | 5.885 | 0.00000063 | 0.00001893 |
| *Unclassified Peptostreptococcaceae_256921.30* | -1.064 | 0.184 | -5.773 | 0.00000234 | 0.00004993 |
| *Facklamia_A_322620 tabacinasalis.8* | 1.803 | 0.225 | 8.005 | 0.00000222 | 0.00004993 |
| *Roseomonas_A_507227 aerofrigidensis.1* | 1.362 | 0.179 | 7.594 | 0.00000250 | 0.00004993 |
| *Lautropia mirabilis.1* | 1.435 | 0.215 | 6.688 | 0.00000284 | 0.00005120 |
| *Moraxella_A_651148 atlantae.1* | 1.172 | 0.198 | 5.911 | 0.00000361 | 0.00005912 |
| *Limosilactobacillus.34* | -0.951 | 0.180 | -5.289 | 0.00002001 | 0.00030020 |
| *Moraxella_C_651924.1* | 0.893 | 0.189 | 4.735 | 0.00003194 | 0.00044220 |
| *Corynebacterium.98* | 1.030 | 0.216 | 4.776 | 0.00003809 | 0.00048973 |
| *Unclassified Enterobacteriaceae_A.1* | 1.630 | 0.286 | 5.690 | 0.00004284 | 0.00051404 |
| *Corynebacterium aquatimens.2* | 0.914 | 0.204 | 4.480 | 0.00005082 | 0.00057175 |
| *Dorea_A longicatena.7* | -0.870 | 0.173 | -5.022 | 0.00006535 | 0.00069199 |
| *Corynebacterium jeikeium.3* | 0.966 | 0.186 | 5.193 | 0.00007333 | 0.00069467 |
| *Unclassified Enterococcaceae.11* | -0.896 | 0.186 | -4.831 | 0.00007102 | 0.00069467 |
| *Paracoccus.112* | -0.964 | 0.172 | -5.591 | 0.00011773 | 0.00100915 |
| *Rothia terrae.8* | 0.793 | 0.174 | 4.559 | 0.00011675 | 0.00100915 |
| *Corynebacterium dentalis.1* | 0.902 | 0.188 | 4.803 | 0.00012361 | 0.00101134 |
| *Kocuria.62* | 0.891 | 0.172 | 5.192 | 0.00017359 | 0.00135857 |
| *Corynebacterium.172* | 0.889 | 0.218 | 4.088 | 0.00020367 | 0.00152754 |
| *Staphylococcus.189* | 0.760 | 0.177 | 4.297 | 0.00024798 | 0.00178549 |
| *Anaerococcus provencensis.13* | 0.899 | 0.180 | 4.986 | 0.00031670 | 0.00211675 |
| *Blastococcus aggregatus.6* | 0.801 | 0.178 | 4.497 | 0.00031751 | 0.00211675 |
| *Micrococcus.19* | 0.772 | 0.203 | 3.810 | 0.00037472 | 0.00240893 |
| *Burkholderia.2* | 1.028 | 0.217 | 4.734 | 0.00039055 | 0.00242411 |
| *Kocuria palustris.9* | 0.856 | 0.222 | 3.865 | 0.00049238 | 0.00295428 |
| *Limosilactobacillus gastricus*.6 | 0.808 | 0.194 | 4.158 | 0.00053383 | 0.00309963 |
| *Deinococcus_B radiophilus*.1 | 0.795 | 0.178 | 4.472 | 0.00076254 | 0.00428927 |
| *Brachybacterium*.46 | 0.661 | 0.175 | 3.770 | 0.00094132 | 0.00513450 |
| *Streptococcus sanguinis_H*.4 | -0.714 | 0.196 | -3.633 | 0.00115880 | 0.00613482 |
| *JC017 sp004296775*.170 | -0.691 | 0.171 | -4.046 | 0.00120296 | 0.00618666 |
| *Corynebacterium*.287 | 0.724 | 0.200 | 3.625 | 0.00149879 | 0.00749395 |
| *Paracoccus sphaerophysae*.1 | 0.620 | 0.180 | 3.445 | 0.00170761 | 0.00823792 |
| *Corynebacterium*.226 | 0.677 | 0.181 | 3.752 | 0.00173912 | 0.00823792 |
| *Romboutsia_B ilealis*.1 | -0.640 | 0.197 | -3.251 | 0.00198409 | 0.00892841 |
| Unclassified *Micrococcaceae*.25 | -0.601 | 0.173 | -3.479 | 0.00194260 | 0.00892841 |
| *Kytococcus sedentarius*.8 | 0.652 | 0.177 | 3.691 | 0.00217872 | 0.00956510 |
| *Moraxella_A_651148 atlantae*.5 | 0.722 | 0.214 | 3.372 | 0.00242934 | 0.01041146 |
| *Paracoccus carotinifaciens*.12 | -0.646 | 0.203 | -3.179 | 0.00250933 | 0.01050415 |
| *Ligilactobacillus ruminis*.2 | -0.644 | 0.188 | -3.416 | 0.00274131 | 0.01096526 |
| *Peptostreptococcus stomatis*.2 | -0.623 | 0.184 | -3.380 | 0.00269893 | 0.01096526 |
| *Dietzia aerolata*.1 | -0.576 | 0.173 | -3.326 | 0.00321258 | 0.01257096 |
| *Staphylococcus*.209 | 0.694 | 0.212 | 3.268 | 0.00337993 | 0.01294443 |
| *Alloprevotella sp900095835*.14 | 0.678 | 0.214 | 3.176 | 0.00394224 | 0.01478340 |
| *Clostridium_T disporicum_203974*.3 | -0.571 | 0.173 | -3.289 | 0.00407796 | 0.01498025 |
| *Kocuria*.32 | 0.639 | 0.196 | 3.256 | 0.00465292 | 0.01675052 |
| *Lactobacillus*.31 | -0.745 | 0.254 | -2.935 | 0.00495771 | 0.01749780 |
| *Weissella_A_338544*.33 | -0.550 | 0.174 | -3.157 | 0.00545081 | 0.01886819 |
| *Marinococcus*.6 | 0.536 | 0.183 | 2.921 | 0.00583946 | 0.01983214 |
| *Corynebacterium glaucum*.10 | -0.566 | 0.183 | -3.087 | 0.00635999 | 0.02119997 |
| *Piscicoccus intestinalis*.4 | -0.537 | 0.188 | -2.859 | 0.00710820 | 0.02284779 |
| *Acinetobacter schindleri*.5 | -0.516 | 0.178 | -2.900 | 0.00705432 | 0.02284779 |
| *Aerococcus*.17 | 0.529 | 0.179 | 2.949 | 0.00741888 | 0.02342805 |
| *Streptococcus anginosus*.3 | -0.570 | 0.179 | -3.194 | 0.00772444 | 0.02397239 |
| *Granulicatella elegans*.11 | 0.638 | 0.233 | 2.742 | 0.00822615 | 0.02467846 |
| *Anaerococcus nagyae*.48 | 0.587 | 0.197 | 2.972 | 0.00816962 | 0.02467846 |
| *Anaerococcus nagyae*.13 | -0.534 | 0.173 | -3.090 | 0.00861713 | 0.02516300 |
| *Paracoccus acridae*.2 | -0.550 | 0.175 | -3.132 | 0.00866726 | 0.02516300 |
| *Peptoniphilus_A*.10 | 0.555 | 0.188 | 2.950 | 0.00994041 | 0.02840116 |
| *Corynebacterium*.201 | 0.624 | 0.239 | 2.607 | 0.01097246 | 0.03086003 |
| *Bacillus_P_294101.36* | 0.516 | 0.175 | 2.943 | 0.01229897 | 0.03304200 |
| *Limosilactobacillus*.40 | -0.544 | 0.195 | -2.789 | 0.01211643 | 0.03304200 |
| *Megasphaera_A_38685 elsdenii.5* | -0.535 | 0.187 | -2.853 | 0.01208127 | 0.03304200 |
| *Kocuria*.29 | 0.485 | 0.176 | 2.751 | 0.01485736 | 0.03932831 |
| *Haemophilus_D_735815.20* | 0.522 | 0.206 | 2.533 | 0.01567157 | 0.04088236 |
| *Staphylococcus*.244 | -0.500 | 0.180 | -2.784 | 0.01652325 | 0.04188992 |
| *Staphylococcus*.304 | 0.485 | 0.188 | 2.576 | 0.01630947 | 0.04188992 |
| *Unclassified Weeksellaceae*.58 | -0.486 | 0.194 | -2.503 | 0.02025225 | 0.05063063 |
| *Coprococcus_A_187866 catus.8* | -0.442 | 0.171 | -2.590 | 0.02139080 | 0.05274443 |
| *Corynebacterium propinquum*.2 | 0.470 | 0.192 | 2.449 | 0.02199119 | 0.05349209 |
| *Catenibacterium*.9 | -0.495 | 0.204 | -2.430 | 0.02262422 | 0.05429814 |
| *Clostridioides_A difficile.5* | -0.445 | 0.192 | -2.322 | 0.02443744 | 0.05787816 |
| *Acinetobacter townneri_A.14* | 0.512 | 0.219 | 2.333 | 0.02478801 | 0.05794601 |
| *Anaerococcus provencensis.7* | 0.483 | 0.199 | 2.430 | 0.02644876 | 0.06103560 |
| *Dolosigranulum pigrum*.19 | -0.525 | 0.229 | -2.293 | 0.02833392 | 0.06455830 |
| *JC017 sp004296775.44* | 0.418 | 0.186 | 2.251 | 0.03239847 | 0.07199660 |
| *Phocaeicola_A_858004 dorei.10* | -0.409 | 0.171 | -2.396 | 0.03232685 | 0.07199660 |
| *Turcibacter*.4 | -0.423 | 0.195 | -2.175 | 0.03307553 | 0.07260483 |
| *Acinetobacter*.205 | -0.404 | 0.184 | -2.194 | 0.03584667 | 0.07773977 |
| *Staphylococcus*.126 | 0.671 | 0.319 | 2.105 | 0.03714380 | 0.07959385 |
| *Unclassified Pseudomonadaceae*.70 | 0.480 | 0.221 | 2.169 | 0.03902092 | 0.08263255 |
| *Rothia kristinae* | -0.614 | 0.290 | -2.120 | 0.03951214 | 0.08269983 |
| *Corynebacterium*.345 | 0.541 | 0.258 | 2.100 | 0.04026508 | 0.08330705 |
| *Metabacillus_B_289979.7* | 0.376 | 0.168 | 2.237 | 0.04088480 | 0.08362801 |
| *Limosilactobacillus fermentum*.5 | 0.457 | 0.215 | 2.124 | 0.04199323 | 0.08398646 |
| *Gemella*.58 | 0.618 | 0.300 | 2.059 | 0.04158316 | 0.08398646 |
| *Halomonas_C_640989.13* | 0.424 | 0.192 | 2.202 | 0.04496234 | 0.08893649 |
| *Acinetobacter*.115 | -0.376 | 0.180 | -2.086 | 0.04555796 | 0.08913514 |
| *Chryseomicrobium excrementi*.2 | 0.380 | 0.177 | 2.143 | 0.04785927 | 0.09263084 |
| *Corynebacterium*.258 | 0.587 | 0.295 | 1.993 | 0.04994428 | 0.09563799 |
| *Agathobacter rectalis*.7 | -0.385 | 0.184 | -2.090 | 0.05535952 | 0.10379909 |
| *Neobacillus*.24 | 0.349 | 0.169 | 2.068 | 0.05525697 | 0.10379909 |
| *Veillonella*_A.12 | 0.603 | 0.316 | 1.910 | 0.05903782 | 0.10955472 |
| *Bacillus*_A.12 | -0.379 | 0.189 | -2.008 | 0.05983546 | 0.10990186 |
| *Rothia* sp001808955.4 | 0.518 | 0.279 | 1.856 | 0.06602250 | 0.12004090 |
| *Unclassified Peptostreptococcaceae*_256921.26 | -0.358 | 0.192 | -1.864 | 0.06741662 | 0.12134992 |
| *Janibacter*_A_390549 *cremeus*.10 | -0.335 | 0.176 | -1.902 | 0.07028515 | 0.12526067 |
| *Clostridium*_T.23 | -0.356 | 0.193 | -1.847 | 0.07120392 | 0.12565398 |
| *Salinicoccus*.28 | -0.425 | 0.242 | -1.762 | 0.08166140 | 0.14270925 |
| *Aeromonas*.1 | 0.338 | 0.192 | 1.767 | 0.08450979 | 0.14626694 |
| *Finegoldia magna*_H.3 | 0.363 | 0.205 | 1.770 | 0.08593940 | 0.14732469 |
| *Veillonella*_A.117 | -0.337 | 0.182 | -1.849 | 0.08738189 | 0.14838435 |
| *Robertmurraya* *korlensis* | 0.316 | 0.173 | 1.829 | 0.09047831 | 0.15079718 |
| *Gracilibacillus dipsosauri* | -0.308 | 0.168 | -1.832 | 0.08995212 | 0.15079718 |
| *Granulicatella*.10 | 0.388 | 0.229 | 1.691 | 0.09640160 | 0.15774807 |
| *Actinomyces*.26 | -0.314 | 0.176 | -1.783 | 0.09623286 | 0.15774807 |
| *Kocuria*.3 | 0.347 | 0.203 | 1.711 | 0.09890108 | 0.15968323 |
| *Faecalibacterium prausnitzii*_C_71358.10 | 0.305 | 0.177 | 1.728 | 0.09935845 | 0.15968323 |
| *Dietzia*.24 | -0.333 | 0.202 | -1.647 | 0.10499603 | 0.16725031 |
| *Lawsonella*.8 | -0.300 | 0.176 | -1.704 | 0.11205783 | 0.17388284 |
| *Microvirga*.111 | -0.292 | 0.169 | -1.722 | 0.11075064 | 0.17388284 |
| *Pauljensenia*.35 | 0.390 | 0.241 | 1.617 | 0.11140033 | 0.17388284 |
| *Dialister* sp900543165.2 | -0.322 | 0.199 | -1.622 | 0.11638803 | 0.17905850 |
| *Veillonella*_A.91 | 0.287 | 0.174 | 1.647 | 0.11896313 | 0.18146919 |
| *Paracoccus*.126 | 0.284 | 0.190 | 1.495 | 0.14358813 | 0.21719213 |
| *Unclassified Enterobacteriaceae*_A.34 | -0.450 | 0.306 | -1.470 | 0.15031972 | 0.22547958 |
| *Pauljensenia* sp000308055.3 | -0.267 | 0.180 | -1.482 | 0.15555388 | 0.23140246 |
| *Alishewanella*_662828.9 | -0.266 | 0.188 | -1.415 | 0.17171268 | 0.25334657 |
| *Corynebacterium kroppenstedtii* | 0.381 | 0.283 | 1.345 | 0.18561375 | 0.27162988 |
| *Chryseobacterium pallidum* | -0.239 | 0.181 | -1.323 | 0.20860711 | 0.30281677 |
| *Anaerostipes hadrus*.10 | -0.212 | 0.172 | -1.232 | 0.23171379 | 0.33366785 |
| *Holdemanella biformis*.18 | -0.222 | 0.178 | -1.251 | 0.23475305 | 0.33536150 |
| *Neisseria*_563205.32 | -0.227 | 0.201 | -1.127 | 0.26648837 | 0.37770005 |
| *Anaerococcus nagsya*.1 | -0.200 | 0.181 | -1.103 | 0.28627887 | 0.40114118 |
| *Streptococcus*.341 | 0.305 | 0.286 | 1.068 | 0.28748451 | 0.40114118 |
| *Unclassified Pasteurellaceae.1* | 0.195 | 0.181 | 1.079 | 0.28996263 | 0.40148673 |
| *Paracoccus aerius.1* | 0.193 | 0.181 | 1.066 | 0.29650022 | 0.40740489 |
| *Kocuria.75* | 0.189 | 0.183 | 1.028 | 0.30748553 | 0.41929845 |
| *Mediterraneibacter_A_155507 faecis.1* | -0.179 | 0.180 | -0.998 | 0.32901468 | 0.44528303 |
| *Corynebacterium.85* | -0.195 | 0.200 | -0.973 | 0.33761351 | 0.45015135 |
| *Ligilactobacillus salivarius.5* | 0.210 | 0.214 | 0.984 | 0.33544171 | 0.45015135 |
| *Bifidobacterium bifidum.1* | 0.211 | 0.226 | 0.932 | 0.35733475 | 0.46608881 |
| *Escherichia_710834.34* | -0.188 | 0.202 | -0.933 | 0.35559328 | 0.46608881 |
| *Corynebacterium durum.2* | -0.168 | 0.174 | -0.964 | 0.35261592 | 0.46608881 |
| *CAG-317 sp000433215.3* | 0.164 | 0.174 | 0.939 | 0.36464930 | 0.47220773 |
| *Unclassified Neisseriaceae_563222.17* | -0.162 | 0.176 | -0.916 | 0.37620768 | 0.48369558 |
| *Streptococcus.38* | -0.238 | 0.277 | -0.857 | 0.39285580 | 0.50151805 |
| *Corynebacterium.32* | 0.170 | 0.199 | 0.857 | 0.39848246 | 0.50511861 |
| *Unclassified Dermatophilaceae_390796.176* | -0.153 | 0.188 | -0.818 | 0.41766463 | 0.52573170 |
| *Unclassified Planococcaceae.34* | -0.136 | 0.172 | -0.794 | 0.43972464 | 0.54965581 |
| *Sarcina ventriculi.12* | -0.136 | 0.178 | -0.765 | 0.45906594 | 0.56987496 |
| *Rothia aerea.1* | -0.128 | 0.171 | -0.747 | 0.46963647 | 0.57900387 |
| *Arthrobacter_D.1* | 0.127 | 0.174 | 0.726 | 0.47639852 | 0.58334512 |
| *Brachybacterium.17* | 0.143 | 0.208 | 0.690 | 0.49293664 | 0.59951754 |
| *Streptococcus.217* | -0.208 | 0.320 | -0.651 | 0.51637905 | 0.62381361 |
| *Unclassified Moraxellaceae.29* | 0.139 | 0.224 | 0.619 | 0.53810741 | 0.64572890 |
| *Acinetobacter baumannii* | -0.123 | 0.210 | -0.585 | 0.56317004 | 0.67132852 |
| *Prevotella copri.25* | 0.116 | 0.203 | 0.568 | 0.57313956 | 0.67871790 |
| *Corynebacterium.118* | 0.087 | 0.172 | 0.507 | 0.61822533 | 0.72732392 |
| *JC017 sp004296775.125* | 0.099 | 0.202 | 0.491 | 0.62646128 | 0.72750342 |
| *Corynebacterium amycolatum_A.1* | 0.105 | 0.212 | 0.494 | 0.62254918 | 0.72750342 |
| *Exiguobacterium_A.10* | 0.083 | 0.218 | 0.381 | 0.70466556 | 0.81307565 |
| *Mesobacillus_A_295502.1* | 0.066 | 0.179 | 0.367 | 0.71608666 | 0.81496691 |
| *Dietzia cinnamea.11* | -0.062 | 0.170 | -0.362 | 0.71988744 | 0.81496691 |
| *Acinetobacter.203* | -0.072 | 0.196 | -0.366 | 0.71846156 | 0.81496691 |
| *Aggregatibacter_736122.3* | -0.057 | 0.180 | -0.314 | 0.75665339 | 0.84798124 |
| *Rothia sp001808955.60* | 0.060 | 0.194 | 0.311 | 0.75847211 | 0.84798124 |
| *Acinetobacter.16* | 0.069 | 0.237 | 0.293 | 0.77040605 | 0.85600673 |
| *Lancefieldella* sp000564995 | 0.059 | 0.217 | 0.273 | 0.78763714 | 0.86978334 |
| *Lactobacillus iners*.6 | 0.052 | 0.201 | 0.258 | 0.79867918 | 0.87422876 |
| *Anaerococcus prevotii*.8 | -0.056 | 0.220 | -0.254 | 0.80137636 | 0.87422876 |
| *Streptococcus infantarius*.3 | 0.051 | 0.220 | 0.230 | 0.81972836 | 0.88886208 |
| Unclassified *Micrococcaceae*.33 | 0.042 | 0.189 | 0.223 | 0.82531210 | 0.88955796 |
| *Neisseria*_563205.31 | 0.037 | 0.197 | 0.189 | 0.85045340 | 0.89521411 |
| *Paracoccus*.78 | -0.037 | 0.185 | -0.198 | 0.84389231 | 0.89521411 |
| *Comamonas*_F_589250.14 | -0.036 | 0.176 | -0.205 | 0.84037966 | 0.89521411 |
| F0422 sp001553345.42 | 0.044 | 0.229 | 0.193 | 0.84813784 | 0.89521411 |
| *Staphylococcus*.119 | 0.029 | 0.167 | 0.176 | 0.86240933 | 0.90252140 |
| *Bifidobacterium*_388775.69 | 0.024 | 0.227 | 0.106 | 0.91619953 | 0.95327119 |
| *Oliverpabstia*.2 | 0.016 | 0.171 | 0.096 | 0.92529662 | 0.95720340 |
| *Anaerococcus*.67 | -0.012 | 0.170 | -0.070 | 0.94533205 | 0.97234153 |
| *Granulicatella elegans*.3 | 0.012 | 0.200 | 0.060 | 0.95299178 | 0.97465068 |
| *Pseudomonas*_A.9 | 0.004 | 0.222 | 0.017 | 0.98660811 | 0.99903696 |
| *Prevotella copri*.31 | 0.003 | 0.206 | 0.015 | 0.98793655 | 0.99903696 |
| *Actinomyces*.14 | 0.000 | 0.230 | -0.001 | 0.99943615 | 0.99943615 |
| *Abiotrophia defectiva*.11 | 0.000 | 0.201 | 0.002 | 0.99815334 | 0.99943615 |

**Table S5.** Results of xgboost models predicting (a) infant birth season and (b) infant growth outcomes from milk microbiota abundances.

| (a) Infant birth season |  |  |  |  |  |
| --- | --- | --- | --- | --- | --- |
| *Study* | *Predicted class* | *AUROC* | *AUPRC* | *Prevalence of rare class* | *p-value* |
| ELICIT | Pre-harvest season | 0.754 | 0.59 | 0.34 | <0.001 |
| ELICIT | Rainy season | 0.554 | 0.52 | 0.49 | 0.15 |
| MISAME-III | Rainy season | 0.759 | 0.41 | 0.21 | <0.001 |
| MUMTA | Rainy season | 0.739 | 0.53 | 0.3 | <0.001 |
| (b) Infant growth outcomes |  |  |  |  |  |
| *Study* | *Predicted class* | *AUROC* | *AUPRC* | *Prevalence of rare class* | *p-value* |
| ELICIT | Stunting at 3 months | 0.686 | 0.44 | 0.19 | 0.003 |
| ELICIT | Stunting at 6 months | 0.461 | -- | -- | -- |
| MISAME-III | Stunting at 3 months | 0.488 | -- | -- | -- |
| MISAME-III | Stunting at 6 months | 0.532 | 0.09 | 0.08 | 0.653 |
| MISAME-III | Small HCAZ at 3 months | 0.582 | 0.05 | 0.6 | -- |
| MISAME-III | Small HCAZ at 6 months | 0.614 | 0.05 | 0.07 | -- |
| MUMTA | Stunting at 3 months | 0.718 | 0.36 | 0.2 | 0.001 |
| MUMTA | Stunting at 6 months | 0.601 | 0.15 | 0.11 | 0.118 |
| MUMTA | Small HCAZ at 3 months | 0.498 | -- | -- | -- |
| MUMTA | Small HCAZ at 6 months | 0.491 | -- | -- | -- |

